# Metabolomics dissection of depression heterogeneity and related cardiometabolic risk

**DOI:** 10.1101/2020.08.11.20172569

**Authors:** Tahani Alshehri, Dennis O Mook-Kanamori, Ko Willems van Dijk, Richard Dinga, Brenda WJH Penninx, Frits R Rosendaal, Saskia le Cessie, Yuri Milaneschi

**Affiliations:** Department of Clinical Epidemiology, Leiden University Medical Center, Leiden, The Netherlands; Department of Public Health and Primary Care, Leiden University Medical Center, Leiden, The Netherlands; Department of Human Genetics, Leiden University Medical Center, Leiden, The Netherlands; Department of Internal Medicine, Division of Endocrinology, Leiden University Medical Center, Leiden, The Netherlands; Donders Institute for Brain, Cognition and Behaviour, Radboud University, Nijmegen, The Netherlands; Department of Psychiatry, Amsterdam Public Health Research Institute, Amsterdam Neuroscience, Amsterdam UMC, Vrije Universiteit, The Netherlands; Department of Biomedical Data sciences, Leiden University Medical Center, Leiden, The Netherlands; GGZ inGeest, Research & Innovation, Amsterdam, The Netherlands

**Keywords:** Metabolomics, Depression, Metabolic Syndrome, Body Fat Distribution, Body Mass Index

## Abstract

**Background:** A recent hypothesis postulates the existence of an “immune-metabolic depression” (IMD) dimension characterized by metabolic dysregulations. Combining data on metabolomics and depressive symptoms, we aimed to identify depressions associated with an increased risk of adverse metabolic alterations.

**Method:** Clustering data were from 1094 individuals with current major depressive disorder and measures of 149 metabolites from a ^1^H-NMR platform and 30 depressive symptoms (IDS-SR30). Canonical correlation analyses (CCA) were used to identify main independent metabolite-symptom axes of variance. Then, for the replication, we examined the association of the identified dimensions with metabolites from the same platform and cardiometabolic endpoints in an independent population-based cohort (n=6572).

**Results:** CCA identified an overall depression dimension and a dimension resembling IMD, in which symptoms such as sleeping too much, increased appetite, and low energy level had higher relative loading. In the independent sample, the overall depression dimension was associated with lower levels of metabolites linked to cardiometabolic risk, such as HOMA-1B −0.09 (95% CI:-0.13--0.06), and visceral adipose tissue −0.15 cm^2^ (95% CI:-0.20--0.10). In contrast, the IMD dimension was associated with well-known adverse cardiometabolic metabolites such as higher visceral adipose tissue 0.11 cm^2^ (95% CI:0.06-0.16), HOMA-1B 0.08 (95% CI: 0.05-0.12), and lower HDL levels −0.04 mmol/L (95% CI:-0.07--0.01).

**Conclusions:** Combining metabolomics and clinical symptoms we identified a replicable depression dimension associated with adverse metabolic alterations, in line with the IMD hypothesis. Patients with IMD may be at higher cardiometabolic risk and may benefit from specific treatment targeting underlying metabolic dysregulations.

## Introduction

Cardiovascular disease (CVD) together with major depressive disorder (MDD) are leading causes of mortality and disease burden worldwide (Dhar & Barton, 2016; Mathers & Loncar, 2006). Each of these conditions may predispose for the other, and the presence of one condition worsens the prognosis of the other (Penninx, Milaneschi, Lamers, & Vogelzangs, 2013). Although the mechanism of this comorbidity is still not fully understood, adverse metabolic alterations may serve as the element that connects the two conditions (Dhar & Barton, 2016; Khandaker et al., 2019; Penninx et al., 2013). A recent large scale epidemiological study in >15,000 individuals analyzing the association between depression and more than 200 lipid related metabolites (Bot et al., 2020) found that depression is associated with a metabolic signature that is also found in CVD patients (Holmes et al., 2018). This metabolic signature was characterized by a shift in the lipids levels encompassing less HDL cholesterol and more very low density lipoproteins (VLDL) and triglycerides, in line with a higher metabolic syndrome profile in depression (Bot et al., 2020). This metabolic signature may represent a substrate linking depression to cardiometabolic diseases (Schwabe et al., 2019). Another large population-based study in >350,000 individuals (Khandaker et al., 2019) concluded that the risk factors of CVD (i.e., inflammatory markers (CRP, IL-6) and biomarker (triglycerides)) are likely causal for the development of depression.

MDD is a highly heterogeneous disorder: patients with the same MDD diagnoses according to DSM-V (Diagnostic and Statistical Manual of Mental Disorders) (American Psychiatric Association, 2013) may experience very different symptom profiles (Lux & Kendler, 2010). These different clinical expressions may be, in turn, differentially related to underlying biological dysregulations. Recent evidence suggests that the MDD link with adverse metabolic alterations and inflammatory dysregulation (i.e., immuno-metabolic dysregulation (Milaneschi, Lamers, Berk, & Penninx, 2020) seems stronger in patients reporting depressive symptoms characterized by altered energy intake/expenditure balance, such as excessive sleepiness, hyperphagia, weight gain, and fatigue (Lamers et al., 2010; Lasserre et al., 2017; Sullivan, Prescott, & Kendler, 2002). Building on this evidence, a recent hypothesis (Milaneschi et al., 2020) postulated the existence of an “immune-metabolic depression” (IMD) dimension emerging from the clustering of energy related clinical symptoms with inflammatory and metabolic dysregulations. Nonetheless, further empirical evidence is needed to fully characterize the clustering between specific symptom profiles and immuno-metabolic biological dysregulations (Fried & Nesse, 2015). The identification of depression dimensions characterized by this clustering of clinical and biological features could give us a better understanding of the shared biological mechanisms between depression and cardiometabolic conditions and potential opening for interventions aimed at avoiding their reciprocal influence (Baune et al., 2012; Fried & Nesse, 2015; Lamers, Milaneschi, de Jonge, Giltay, & Penninx, 2018). Furthermore, the identification of individuals with this specific form of depression may create awareness amongst healthcare providers and the need to perform more rigorous cardiometabolic health checks and interventions.

The main aim of the present study was to identify depression dimensions associated with increased risk of adverse metabolic profile by combining data on metabolomics and depressive symptoms. First, we applied a data-driven method to identify patterns of correlations between depressive symptoms and metabolites from a lipid-focused metabolomic platform in >1,000 MDD patients. Then, for the replication, we examined the association between the identified dimensions and 51 metabolites from the same panel, and clinical cardiometabolic biomarkers such as fasting glucose, insulin resistance, total and abdominal adiposity in an independent population-based cohort (n=6572).

## Method

### Study design

The current analysis consists of two parts: the metabolite-symptom clustering and the replication (**Figure 1**). In the first part, we used a data-driven approach to dissect the heterogeneity of depression and to identify main independent metabolite-symptom dimension of variance in 1094 individuals with current (i.e., within the past 6 months) depression from the Netherlands Study of Depression and Anxiety cohort (NESDA). Then, in the replication, we examined the association between the dimensions identified and the cardiometabolic metabolites (51 lipids, fatty acids, and low-molecular-weight metabolites) and endpoints in an independent dataset of 6572 participants from the general population enrolled in the Netherlands Epidemiology of Obesity (NEO) study. The research protocol of NESDA was approved by the medical ethical committees of the following participating universities: Leiden University Medical Center (LUMC), Vrije University Medical Center (VUMC), and University Medical Center Groningen (UMCG). The NEO study was approved by medical ethics committee of Leiden University Medical Center (LUMC). All participants gave written informed consent.

**Figure 1.**
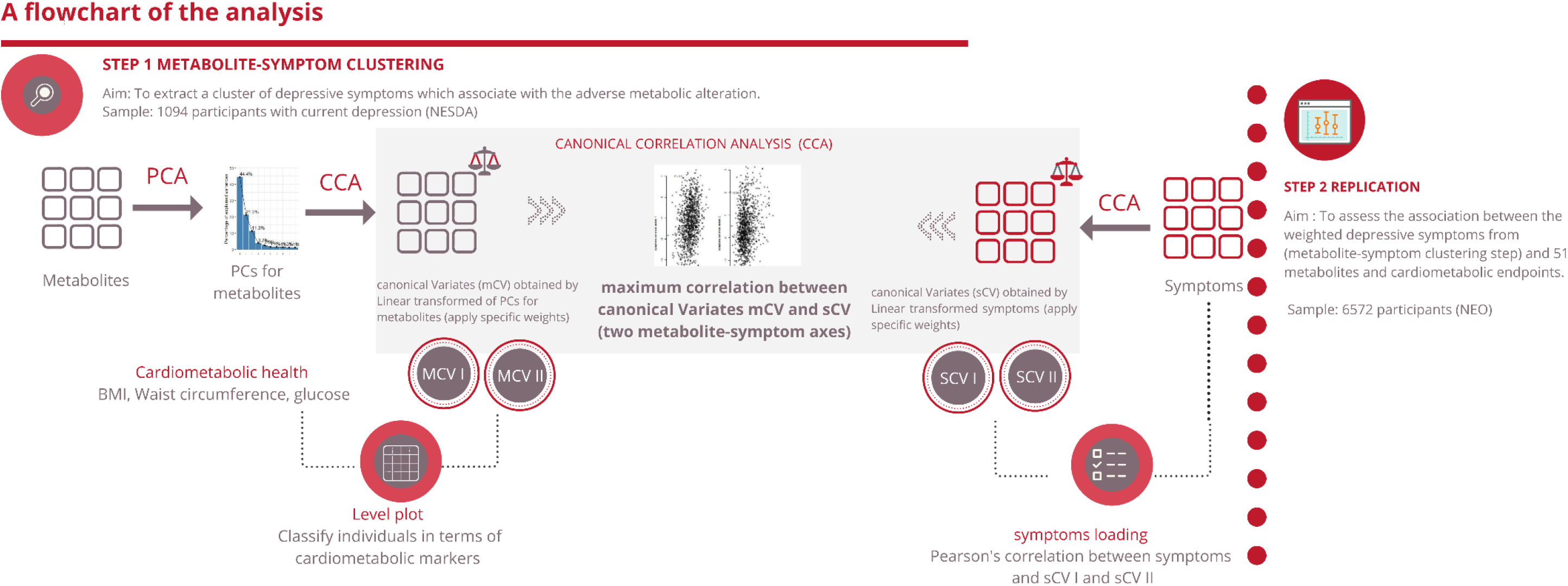
An illustration of the method

### Part 1: Metabolite-symptom clustering

We performed this analysis on 1094 participants diagnosed with current (i.e., within the past 6 months) MDD via the structured Composite Interview Diagnostic Instrument (CIDI, version 2.1) (Robins et al., 1988) from NESDA (Penninx et al., 2008). After an overnight fast, EDTA plasma was collected and stored in aliquots at −80°C until further analysis by ^1^H-NMR Nightingale Health Ltd, Helsinki, Finland (Soininen, Kangas, Wurtz, Suna, & Ala-Korpela, 2015) metabolomics platform. This metabolomics platform consists of 230 metabolites or metabolite ratios and can be classified into 3 clusters (Wurtz et al., 2017) as follow: 1) lipids, fatty acids, and low-molecular-weight metabolites (n= 51); 2) lipid composition and particle concentration measures of lipoprotein subclasses (n= 98); and 3) metabolite ratios (n= 81). In this analysis, we focused on the first two classes (n=149). Metabolite ratios were not used due to redundancy. Values of metabolites that could not be quantified were set as missing for all individuals. Furthermore, metabolite values with outlying concentrations (± 5 SD) were additionally set as missing. A value of 1 was added to all metabolite values, which were subsequently natural log-transformed to approximate normality. The obtained values were scaled to standard deviation units to enable comparison. This protocol for processing the metabolomic data was suggested by the manufacturer of the platform and has been consistently applied in several large-scale epidemiological studies (Bot et al., 2020; Onderwater et al., 2019). Blood samples were analyzed in two batches (April 2014 and December 2014) by ^1^H-NMR Nightingale Health Ltd, Helsinki, Finland) (Soininen et al., 2015). A variable indexing batches was added as covariate in the analysis in order to account for potential batch effect. We regressed the metabolites on age and batch effect in order to remove their confounding effect.

During the baseline assessment, the presence of major depressive disorder was determined with the Diagnostic and Statistical Manual of Mental Disorders, Fourth Edition (DSM-IV)-based Composite Interview Diagnostic Instrument (CIDI, version 2.1, World Health Organization, 1997) by specially trained research staff. The CIDI has a high reliability and validity for assessment of depressive disorders (Wittchen, 1994). In addition, participants were asked to complete a Dutch translation of the Inventory of Depressive Symptomatology (IDS-SR30), which assesses (via a 4-level response system) the presence of 30 depressive symptoms during the last week and their severity (Rush, Gullion, Basco, Jarrett, & Trivedi, 1996).

Body mass index (BMI), waist circumference and fasting glucose level were also used in the analysis to examine the relationship between CCA output and cardiometabolic health. Height and weight were measured to calculate BMI in kg/m^2^ as an index of general adiposity. Waist circumference (cm), defined as the minimal abdominal circumference between the lower edge of the rib cage and the iliac crests, was measured by trained clinical staff according to a standardized procedure as index of abdominal adiposity. Glucose was measured from fasting plasma samples by using standard laboratory technique.

#### Statistical analysis for metabolite-symptom clustering

Our goal was to identify independent dimensions emerging from patterns of correlations between depressive symptoms and metabolites. To do this, we used canonical correlation analysis (CCA, (Hotelling, 1936)). CCA is a method that given two sets of variables X and Y (in this case, metabolites and depressive symptoms), find a linear combination of X that is maximally correlated with a linear combination of Y (i.e., a weighted sum of each variable). The linear transformation weights were chosen such that the correlation between resulting linear combinations is maximized. These linear combinations are called canonical variates (i.e., mCV (metabolites canonical variates), sCV (symptoms canonical variates)). Together mCV and sCV are called a canonical pair and the correlation between this canonical pair is called the canonical correlation. In a specific dataset, it is possible to find multiple canonical pairs such that canonical pairs are uncorrelated to each other and equal to the number of variables in the smallest dataset. In our analysis we chose to proceed with the first two canonical pairs that provided more information about the two sets of variables. The relationship between the created canonical variables of depressive symptoms and metabolites from the same panel and cardiometabolic endpoints was validated in an independent sample (see replication section).

Because metabolites are highly correlated to each other, we first reduced the 149 metabolites residuals into 3 variables that explained 75% of variance in the data using principal component analysis (PCA). PCA is a method that transforms the variables so they maximize the variance in the data, from which we selected components with the highest variance (David & Jacobs, 2014). Therefore, the next analysis was performed on metabolic variables explained the highest variance (principal components (PC)) and 30 depressive symptoms.

In order to explore how the first two metabolic canonical variates (mCVI and mCVII) classify individuals in terms of conventional cardiometabolic biomarkers (i.e., BMI, waist circumference, fasting glucose) we plotted the predicted level of the biomarkers as a function of the two metabolic canonical variates (i.e., smoothing function was used for the prediction). After that, to evaluate the symptoms contribution to the two canonical correlation, for each symptom we calculated the symptoms loadings, expressed in Pearson’s correlation coefficient, with the first two symptoms canonical variates (sCVI and sCVII).

### Part 2: Replication

To replicate the results of previous step, we investigated the association between the dimensions identified in the previous step via CCA and metabolomics and clinical endpoints in the Netherlands Epidemiology of Obesity (NEO) study (de Mutsert et al., 2013). The depressive symptoms in NEO study were assessed by IDS-SR30 (Rush et al., 1996), the same instrument used in the NESDA study. For the purpose of replication, we included only the first class from the ^1^H-NMR platform (i.e., 51 lipids, fatty acids, and low-molecular-weight metabolites) in the main results. For completeness of data, we showed the result of the entire metabolomic platform in the supplementary results since they have large overlap with the standard clinical lipid profile. We used the same protocol for processing this metabolomic data in the clustering step. The cardiometabolic endpoints are described in detail elsewhere (de Mutsert et al., 2013). From fasting glucose and insulin concentrations, we calculated the Homeostasis Model Assessment for Insulin Resistance (HOMA-IR) and HOMA of beta-cell function (HOMA-1B) as markers of hepatic insulin resistance and steady-state insulin secretion (Matthews et al., 1985). HOMA-IR was calculated as fasting insulin (μU/mL) x fasting glucose (mmol/L)/22.5 and HOMA-1B% as 20 x fasting glucose (mmol/l)-3.5 (Matthews et al., 1985; Wallace, Levy, & Matthews, 2004).

#### Statistical analysis for replication

To index the two dimensions identified in the clustering step, we created two weighted depressive symptom scores. We weighted each individual item of the IDS-SR30 based on extracted CCA weights from the previous step. Then, we summed the weighted depressive symptoms to create two weighted IDS scores. We used linear regression to examine the relationship between the two weighted IDS scores as the independent variable and 51 ^1^H-NMR metabolites and cardiometabolic endpoints (BMI, total body fat, waist circumference, visceral adipose tissue, HbA1c, fasting glucose, HOMA-IR, HOMA-1B, total cholesterol, LDL-cholesterol, HDL-cholesterol, and triglycerides) as dependent variables. We fitted four linear regression models, the crude model, model 1, model 2 and, model 3. Model 1 was adjusted for age, sex, and educational level. Model 2 was adjusted for age, sex, educational level, smoking, alcohol consumption, physical activity, and ethnicity. Model 3 was model 2 with additional adjustment for lipid-lowering drugs, and antidepressants. In the investigation of the association between weighted depressive symptoms score and the 51 ^1^H-NMR metabolites, the false discovery rate (FDR) method was applied to correct for the multiple testing. As the NEO study is a population-based study with oversampling of individuals with a BMI > 27 kg/m^2^, all results are based on weighted analysis toward BMI distribution in the general Dutch population.

## Results

### Metabolite-Symptom clustering

Table 1 shows the main demographic, health- and depression-related characteristic, in the NESDA sample of individuals with current MDD. Data reduction of metabolomics was performed using PCA, identifying three principle components that explained together 75% of the variance. The resulting 3 principle components were used in the CCA analysis and were correlated to the 30 depressive symptoms, to identify the main independent metabolite-symptom dimensions of variance based on their correlation. The correlation between the linear transformation (weights) of metabolites principal components (metabolic canonical variate I, mCVI) and depressive symptoms (symptom canonical variate I, sCVI) was 0.30 explaining 54 % of the metabolite-symptom covariance, for the second pair of canonical variates the correlation between mCVII and sCVII was 0.24 explaining 32% of the metabolite-symptom covariance (Supplemental figure 2).

**Table 1.**
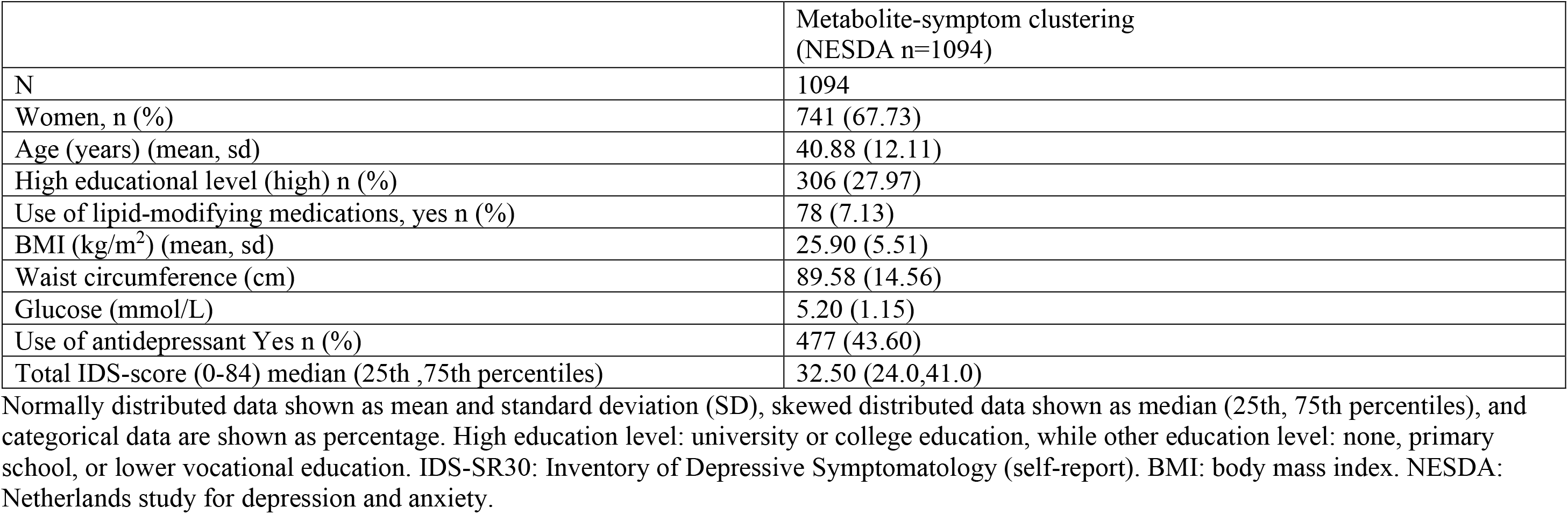
Characteristics of the study population for the metabolite-symptom clustering step (NESDA)

|  | Metabolite-symptom clustering<br>(NESDA n=1094) |
| --- | --- |
| N | 1094 |
| Women, n (%) | 741 (67.73) |
| Age (years) (mean, sd) | 40.88 (12.11) |
| High educational level (high) n (%) | 306 (27.97) |
| Use of lipid-modifying medications, yes n (%) | 78 (7.13) |
| BMI (kg/m<sup>2</sup>) (mean, sd) | 25.90 (5.51) |
| Waist circumference (cm) | 89.58 (14.56) |
| Glucose (mmol/L) | 5.20 (1.15) |
| Use of antidepressant Yes n (%) | 477 (43.60) |
| Total IDS-score (0-84) median (25th ,75th percentiles) | 32.50 (24.0,41.0) |

Normally distributed data shown as mean and standard deviation (SD), skewed distributed data shown as median (25th, 75th percentiles), and categorical data are shown as percentage. High education level: university or college education, while other education level: none, primary school, or lower vocational education. IDS-SR30: Inventory of Depressive Symptomatology (self-report). BMI: body mass index. NESDA: Netherlands study for depression and anxiety.

In order to explore how the first two metabolic canonical variates (mCVI and mCVII) classify individuals in terms of conventional cardiometabolic biomarkers (i.e., BMI, waist circumference, fasting glucose) we plotted the predicted level of the biomarkers as a function of the two metabolic canonical variates. Level plots depicted in Figure 2 show that high values in BMI, waist circumference, and fasting glucose tended to cluster at high level of mCVII and low levels for mCVI. Figure 3 shows the loading, expressed as Pearson’s correlation coefficient, of IDS-SR item on the two symptoms canonical variates (sCVI and sCVII). In the first variate, correlation coefficients were substantially consistent across the entire spectrum of items, including mood, cognitive and somatic symptoms. In the second variate, the loading of specific items such as difficulties falling asleep, sleeping too much, increase weight and appetite, low energy level and gastrointestinal problems were relatively higher as compared to the other symptoms.

**Figure 2.**
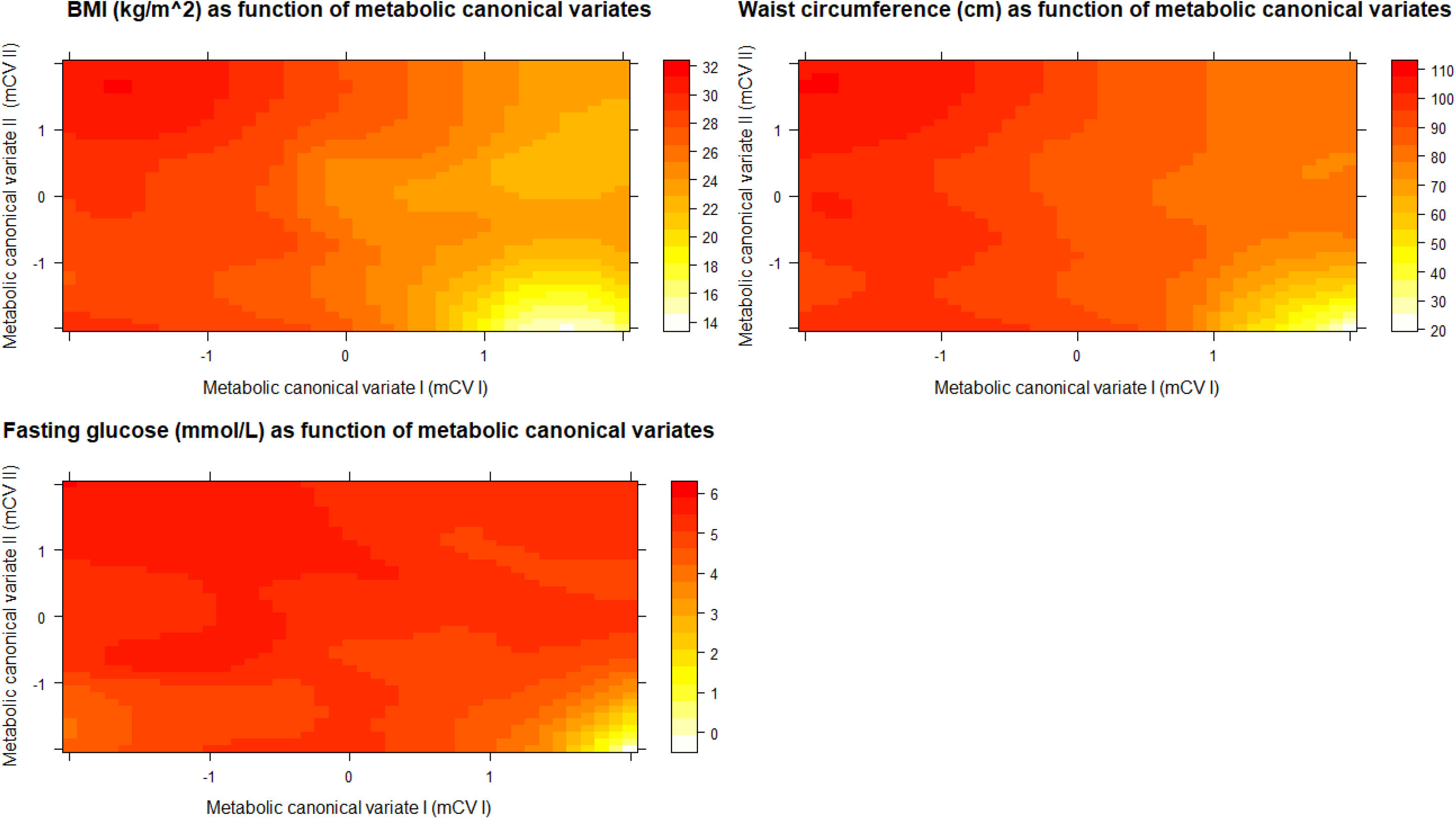
Level plot of the predicted cardiometabolic health conventional biomarker as functions of the first and second metabolic canonical variates.

**Figure 3.**
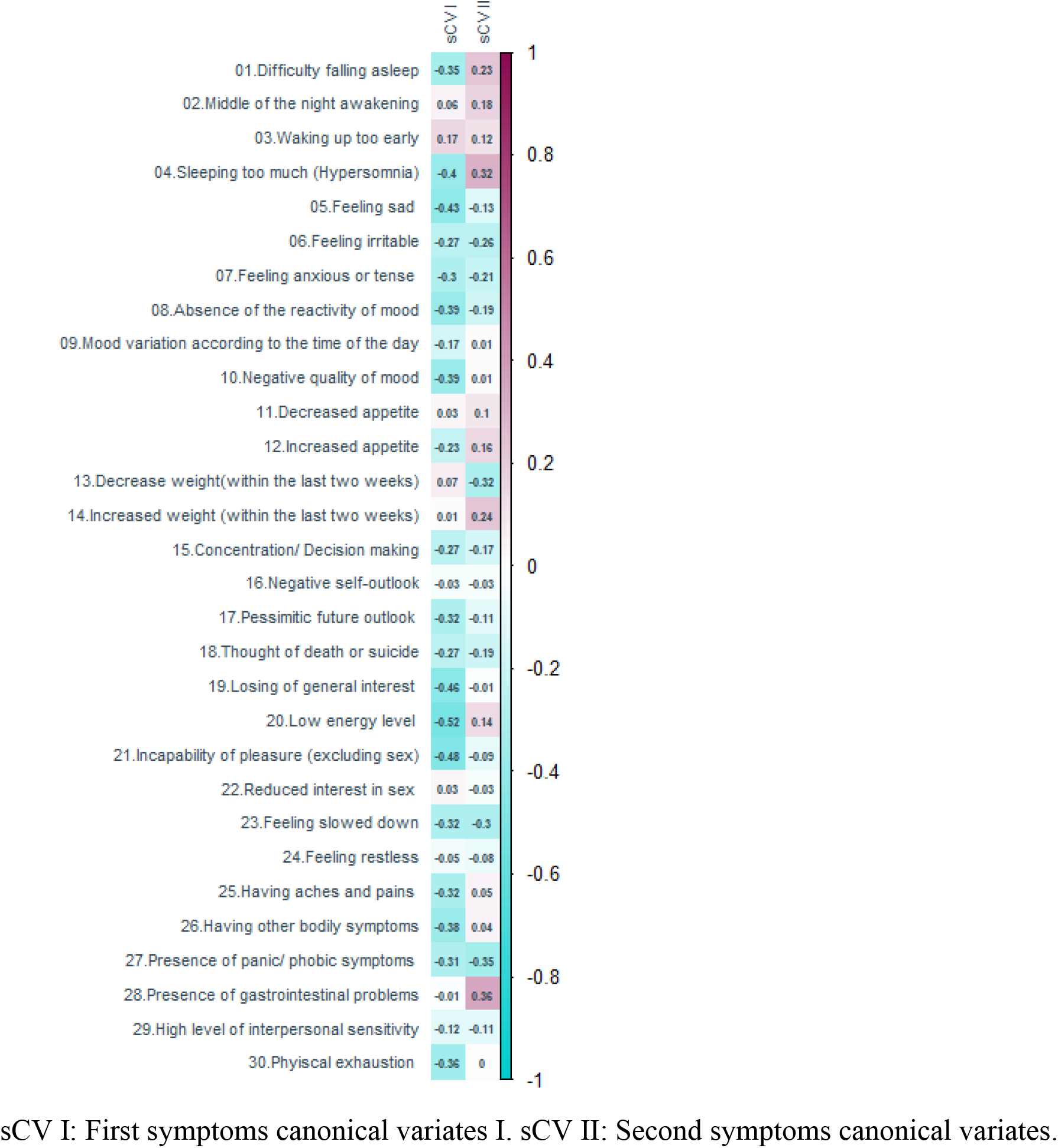
Canonical loading of depressive symptoms on the symptoms canonical variates

We interpreted the first canonical variate CVI, explaining a larger proportion of symptom-metabolite covariance (54%), as an overall depression dimension characterized by a wide array of symptoms (sCVI, Figure 3) and lower levels of cardiometabolic biomarkers (mCVI, Figure 2). The second variate, explaining 32% of the symptom-metabolite covariance, partially resembled the postulated IMD construct (Milaneschi et al., 2020), with relevance for energy-related behavioral symptoms and higher cardiometabolic biomarkers. Thus, for interpretability we labelled the two canonical variates, respectively, “overall depression” and “IMD”.

### Replication

The baseline characteristics for all 6572 participants of the NEO cohort included in the replication step are shown in Supplemental table 1. The mean age in the NEO population was 55.7 years (standard deviation (SD)): 6 years, and the median of the IDS-SR30 questionnaire was 8.0 points (4, 13). We created two weighted depressive symptoms scores labelled “overall depression” and “IMD” with the weights derived in CCA for, respectively, the first and second canonical variate. We examined the association of these weighted scores with 51 metabolites and cardiometabolic endpoints (BMI, total body fat, waist circumference, visceral adipose tissue, HbA1c, fasting glucose, HOMA-IR, HOMA-1B, total cholesterol, LDL-cholesterol, HDL-cholesterol and triglycerides). Figures 4A and 4B depict the linear regression effect estimates and 95% confidence intervals for the association between the weighted symptom sum score and the 51 lipids, fatty acids, and low-molecular-weight metabolites, and cardiometabolic endpoints adjusted for age, sex, and educational level (model 1). Results of all crude and adjusted models can be found in Supplemental table 2 and 3. In general the two weighted symptoms scores showed divergent pattern of results: IMD showed metabolic alterations linked to increased cardiometabolic risk, while overall depression score showed opposite associations. IMD was associated with (per standard deviation) higher glycoprotein acetylase 0.12 mmol/L (95% CI: 0.08-0.15), apolipoprotein B 0.08 g/L (95% CI:-0.05-0.12), triglyceride levels 0.12 mmol/L (95% CI: 0.09-0.15), total body fat 0.09% (95% CI:0.06-0.11), visceral adipose tissue 0.11 cm^2^ (95% CI:0.06-0.16), HOMA-1B 0.08 (95% CI: 0.05-0.12), and lower HDL levels −0.04 mmol/L (95% CI: −0.07-0.01). In contrast, the overall depression was associated with (per standard deviation) glycoprotein acetylase −0.16 mmol/L (95% CI: −0.20--0.13), apolipoprotein B −0.06 g/L (95% CI:-0.09--0.02), triglyceride levels −0.12 mmol/L (95% CI: −0.16--0.09), total body fat - 0.11% (95% CI:-0.13--0.09), visceral adipose tissue −0.15 cm^2^ (95% CI:-0.20—0.10), HOMA-1B −0.09 (95% CI:-0.13--0.06), and HDL levels 0.10 mmol/L (95% CI: 0.07-0.13) (Figure 4A, 4B). We repeated the analysis of the linear regression with additional adjustment for lipid lowering drugs (model 3) and results did not notably change (Supplement table 2,3)

**Figure 4.**
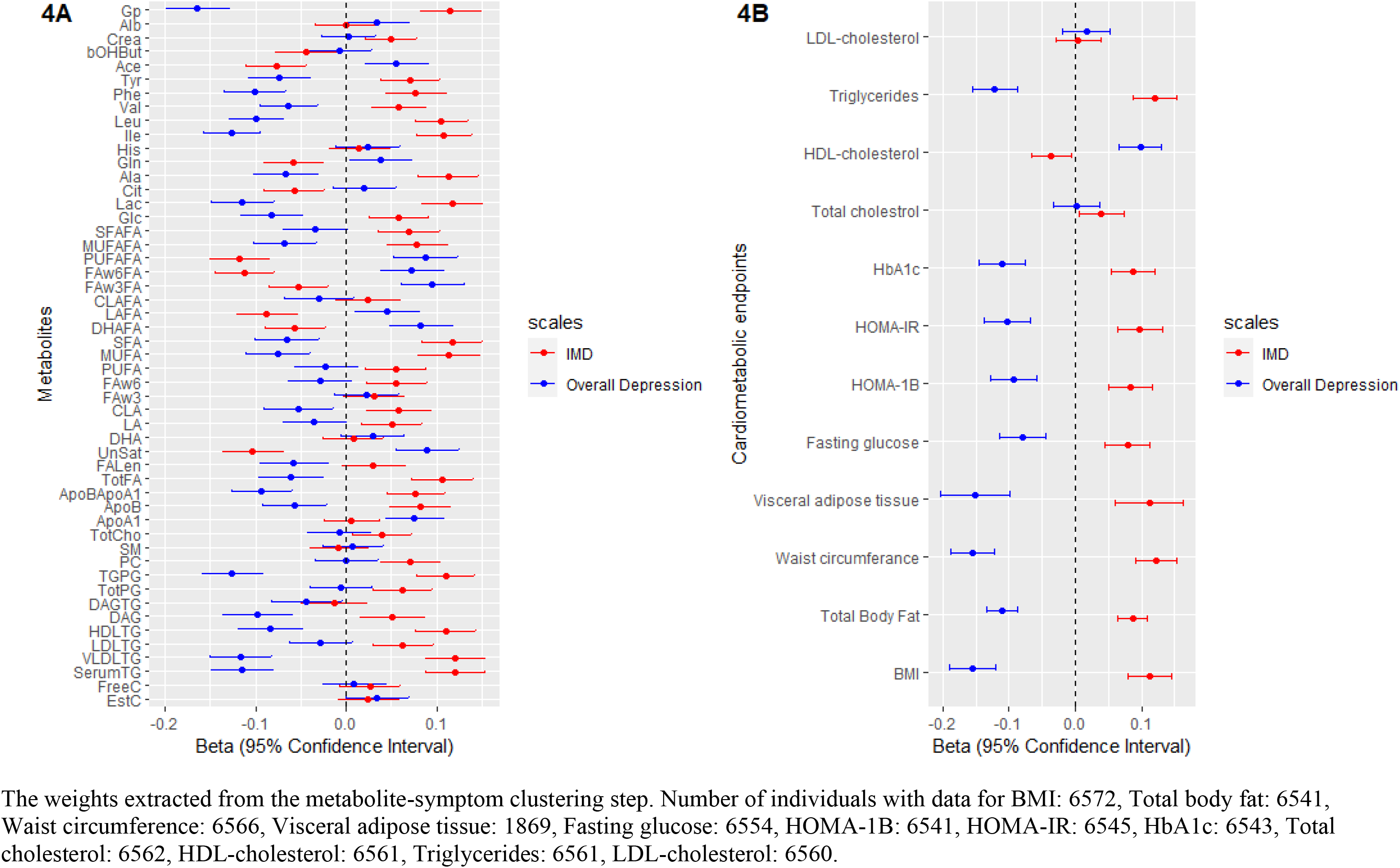
The linear regression analysis of the association between the weighted depressive symptoms score and the cardiometabolic endpoints and metabolites in individuals from NEO study.

## Discussion

Using a data-driven method, we combined metabolomics and clinical symptoms data to dissect depression heterogeneity and identify independent underlying dimensions in participants diagnosed with current MDD from NESDA cohort (n=1094). Then, we replicated our results by examining the association between the identified dimensions and 51^1^ metabolites from the same lipidomic panel, and cardiometabolic endpoints in an independent dataset of 6572 participants from the general population enrolled in the Netherlands Epidemiology of Obesity (NEO) study.

We identified a major dimension reflecting overall depression explaining a large proportion (54%) of symptom-metabolite covariance, and characterized by a wide array of symptoms and reduced levels of cardiometabolic biomarkers. A second dimension explaining 34% of symptom-metabolite covariance emerged as characterized by higher cardiometabolic biomarkers and higher relative relevance for symptoms such as difficulties falling asleep, sleeping too much, increase weight and appetite, low energy level and gastrointestinal problems. This second dimension partially resemble the recently pustulated (Milaneschi et al., 2020) construct of IMD, defined by the clustering of inflammatory and metabolic dysregulations with behavioral energy-related symptom. We labelled therefore the first and second dimensions “overall depression” and “IMD”. In the replication step, we found that the IMD dimension was associated with a metabolic profile similar to the metabolic profile reported in individuals with high cardiometabolic risk profile such as higher triglyceride levels, lower HDL-cholesterol levels, higher visceral adipose tissue, higher branched chain amino acids, and higher glycoprotein acetylase and insulin resistance. In contrast, the associations between these metabolites and the overall depression dimension were in the opposite direction, indicating a lower cardiometabolic risk.

The present finding confirm the presence of partially divergent correlation structures between specific depressive symptom profiles and metabolic dysregulations. This is in line with the previous research in this field that confirmed that the presence of homeostatic shift toward increase energy (increased appetite) intake and decrease energy expenditure (sleeping too much, difficulty falling asleep (Markwald et al., 2013) and low energy level) were more strongly associated with inflammatory and metabolic biomarkers considered as risk factors for CVD. In earlier work based on NESDA data, among participants with active depression episode, increased a neuroendocrine energy homeostasis marker (leptin) (Zakrzewska, Cusin, Sainsbury, Rohner-Jeanrenaud, & Jeanrenaud, 1997) was associated (independently from BMI) with a depressive symptoms profile defined by increase the intake (increase appetite/weight) and decrease the expenditure (fatigue, low energy) (Milaneschi, Lamers, Bot, Drent, & Penninx, 2017). Likewise, in the same population, another study confirmed the relationship between cardiometabolic conditions, such as abdominal adiposity, inflammation markers, and metabolic syndrome, and increased appetite during the active depressive episode (Lamers et al., 2018). In agreement with above-mentioned well characterized clinical cohort studies, similar results were obtained from two large population-based studies (Alshehri et al., 2019; Jokela, Virtanen, Batty, & Kivimaki, 2016) that confirmed the association between this cluster of symptoms and higher CRP and adiposity. Also, in a small study that combined neuroimaging and biochemical approaches, hyperphagia during depression was strongly associated with endocrine dysregulation and inflammation (Simmons et al., 2016). Interestingly, earlier (Yuri Milaneschi et al., 2017) and recent (Badini et al., 2020) genomic studies in > 25,000 and > 30,000 found that the genetic overlap between BMI, CRP and leptin with depression is symptom specific; this overlap was only found in depressed patients with increased weight and appetite. Another study (Adams et al., 2019) that used neuroticism as genetic specifier to stratify depression patients showed that the portion of the common genetic liability between depression and neuroticism was also share with other psychiatric disorders; interestingly, the genetic liability not shared with neuroticism was positively correlated with metabolic phenotypes and cardiovascular disease. These results confirm the existence of different dimension within the construct of depression rooted in underlying biological and genetic mechanisms. Based on evidence along this line of research, the existence of an “immuno-metabolic depression (IMD)” dimension of depression was hypothesized (Milaneschi et al., 2020). This dimension is characterized by the clustering of immuno-metabolic biological alterations and behavioral symptom related to homeostasis dysregulation, which in turn can be the link between depression and cardiovascular disease (Milaneschi et al., 2020).

Many plausible mechanisms can directly or indirectly lead to or result from this homeostatic shift as maintaining energy homeostasis is governed by biological, behavioral and environmental factors (Chapelot & Charlot, 2019). For example (Milaneschi et al., 2020), low-grade inflammation which associated with adiposity and depression (Woelfer, Kasties, Kahlfuss, & Walter, 2019), favor -as proposed previously (Lacourt, Vichaya, Chiu, Dantzer, & Heijnen, 2018)- the fast aerobic glycolysis in the immune cells over other efficient but yet slower energy production pathways (e.g., lipid oxidation). This appropriation of the available cellular fuel done by immune cells results in low energy available to any other activities. When the body has low energy level, the circadian rhythm and sleep cycle disturb as well (i.e., feeling tired and sleeping during the day which affect sleeping time and quality during the night) (Lacourt et al., 2018). Moreover, dysregulation of neuroendocrinological signaling such as leptin, and insulin (which have crucial metabolic roles) may diminish their function as satiety inducers hormones which lead to the development of increased appetite and decreased energy level symptoms (Chapelot & Charlot, 2019). These biological processes also interact with behavioral/environmental factors that contribute in regulating of the energy homeostasis. Obesogenic environment (e.g., low physical activity demand, and availability of palatable food) could shift the energy balance toward energy accumulation which in turn can result in low grade inflammation and neuroendocrinal dysregulation (Church & Martin, 2018; Thyfault, Du, Kraus, Levine, & Booth, 2015). Putting it together, this symptoms profile (difficulty falling asleep, sleeping too much, increased appetite, low energy level) may reflects a prolonged homeostatic failure that closely interconnected with neuroendocrinal and metabolic dysregulation that also reported in patients with CVD from metabolic dysregulation (Naisberg, 1996).

Fully characterizing the IMD dimension identified in the present study, in terms of its clinical manifestation and underlying biological mechanisms is the first step in the path to a personalized approach for patients with depression (Simon & Perlis, 2010). This full characterization may help in guiding the choice of the most suitable intervention to alleviate the symptoms burden or to prevent its adverse prognosis. Moreover, understanding the clinical, and biological characteristics of this depression dimension will increase the precision of the genetic studies that aim to comprehend depression genetic architecture (Schwabe et al., 2019). Future research is needed to help us understand to what extent treating underlaying metabolic dysregulation (i.e., lipids and glucose) will contribute to mitigate this symptoms profile adversity. On the other hand, we also need to know to what degree will behavioral intervein that target this symptoms profile such as exercising, dieting and sleep hygiene will play a role in achieving better cardiometabolic health profile. Moreover, future genetics studies using techniques such as Mendelian Randomization are needed to test the causal direction between metabolic dysregulation (i.e., triglycerides (Khandaker et al., 2019)) and specific depressive symptom profile.

To the best of our knowledge, this study is the largest study that exploits jointly metabolomic data to dissect the dimensionality of depressive symptoms in a large, well-defined clinical cohort (NESDA). Moreover, we replicate our findings from the clustering set in a population based large cohort (NEO). However, some methodological issues should be considered. First, we should acknowledge the limitation of the NMR metabolite platform, which mainly is a lipidomic metabolomic platform. For that reason, the term metabolic dysregulation should be interpreted based on the used metabolomic platform (i.e., metabolic dysregulation in this lipid focus metabolomic platform). Clustering based on different metabolomic platform could results on divergent results. However, the metabolomic platform used in this study is sufficient for inspecting the association between depression symptomatology and cardiometabolic endpoints (Holmes et al., 2018). Third, based on the cross-sectional study design, we are unable to infer the directionality of the relationship between depressive symptoms and adverse metabolic alterations.

In the present study, using a data-driven method we identified two independent depression dimension differentially related with cardiometabolic risk markers, such as higher triglycerides, higher visceral fat content, lower HDL-cholesterol levels and insulin resistance in the replication step. Our findings confirm that depression is associated with metabolic alterations that could represent the mechanism linking depression with cardiometabolic disorders. However, these metabolic alteration are not present in all forms of depression. Depressed patients with IMD may be at higher cardiometabolic risk and may require specific additional treatment targeting underlying metabolic dysregulations.

## Data Availability

Data are not publicly available

## Acknowledgement

**NESDA**

The infrastructure for the NESDA study (**www.nesda.nl**) is funded through the Geestkracht program of the Netherlands Organization for Health Research and Development (ZonMw, grant number: 10-000-1002) and financial contributions by participating universities and mental health care organizations (VU University Medical Center, GGZ inGeest, Leiden University Medical Center, Leiden University, GGZ Rivierduinen, University Medical Center Groningen, University of Groningen, Lentis, GGZ Friesland, GGZ Drenthe, Rob Giel Onderzoekscentrum).

**NEO study**

The NEO study is supported by the participating Departments, Division, and Board of Directors of the Leiden University Medical Center, and by the Leiden University, Research Profile Area Vascular and Regenerative Medicine. DOM-K is supported by Dutch Science Organization (ZonMW-VENI Grant No. 916.14.023).We express our gratitude to all participants of the Netherlands Epidemiology of Obesity (NEO) study, in addition to all participating general practitioners. We furthermore thank P.R. van Beelen and all research nurses for collecting the data, P.J. Noordijk and her team for sample handling and storage, and I. de Jonge for data management of the NEO study.

## Disclosures of conflicts of interest

Dennis Mook-Kanamori reports personal fees from Metabolon, Inc, outside the submitted work, and Brenda Penninx reports grants from Jansen Research and Development, LLC, and grants from Boehringer Ingelheim, outside the submitted work. All other co-authors declared no conflict of interest.

---

**Supplemental table 1.** Characteristics of the replication step, the Netherlands epidemiology of obesity (NEO) study

|  | Replication |
| --- | --- |
| Women (%) | 56.34 |
| Age (years) (mean, sd) | 55.70 (6.0) |
| High educational level (high) (%) | 46.08 |
| Tobacco smoking n% |  |
| Never (%) | 38.68 |
| Former (%) | 45.25 |
| Current (%) | 16.07 |
| Alcohol consumption (g/day) median | 9.86 (2.74,21.41) |
| Use of lipid-modifying medications, yes (%) | 10.31 |
| BMI (kg/m<sup>2</sup>) (mean, sd) | 26.32(4.43) |
| Total body fat (%) | 31.6 (24.8,38.2) |
| Waist circumference (cm) | 92.15 (13.34) |
| Visceral adipose tissue (cm<sup>2</sup>) | 90.04 (56.04) |
| Serum concentration of total cholesterol (mmol/L) | 5.68 (1.05) |
| Serum concentration of LDL (mmol/L) | 3.52 (0.96) |
| Serum concentration of HDL (mmol/L) | 1.57 (0.46) |
| Serum concentration of triglyceride (mmol/L) | 1.23 (0.84) |
| HBA1C (%) | 5.36 (0.46) |
| Glucose (mmol/L) | 5.47 (0.96) |
| HOMA-IR | 1.86 (1.21,2.93) |
| HOMA 1B | 86.67 (60.0,126.00) |
| Use of antidepressant Yes (%) | 6.59 |
| Total IDS-score (0-84) median (25th, 75th percentiles) | 8 (4,13) |

Number of individuals with data for BMI: 6572, Total body fat: 6541, Waist circumference: 6566, Visceral adipose tissue: 1869, Fasting glucose: 6554, HOMA-1B: 6541, HOMA-IR: 6545, HbA1c: 6543, Total cholesterol: 6562, HDL-cholesterol: 6561, Triglycerides: 6561, LDL-cholesterol: 6560. Normally distributed data shown as mean and standard deviation (SD), skewed distributed data shown as median (25th, 75th percentiles) and categorical data are shown as percentage. High education level: university or college education, while other education level: none, primary school or lower vocational education. IDS-SR30: Inventory of Depressive Symptomatology (self-report). BMI: body mass index. HOMA-IR: (homeostatic model assessment insulin resistance)= (glucose x insulin)/22.5, HOMA-1B: (homeostatic model assessment of beta cells) $HOMA\ 1B = 20 \times \text{insulin} / \text{glucose} - 3.5$

**Supplemental figure 1:**
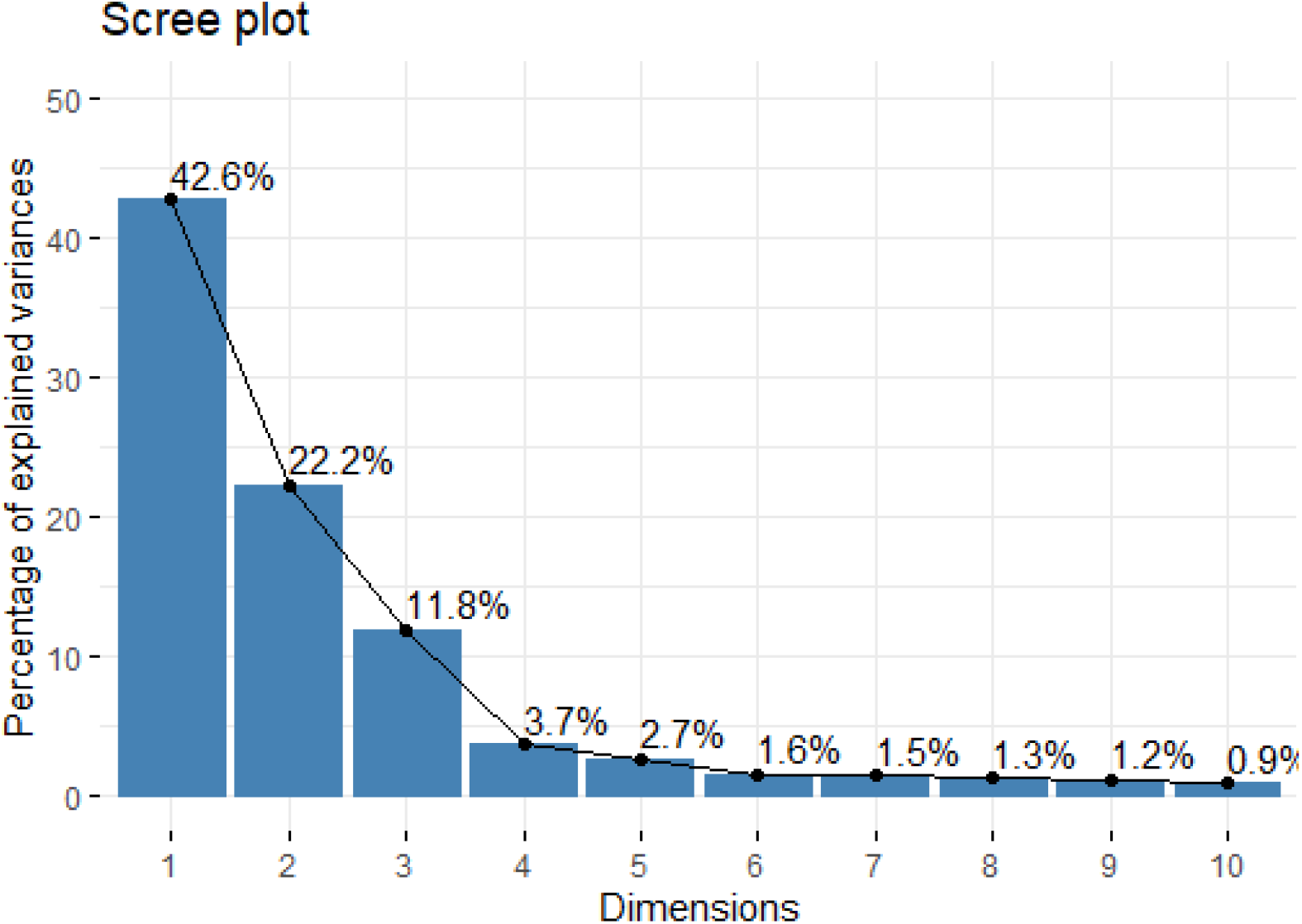
Scree plot for the principle component analysis of the metabolites dataset.

**Supplemental figure 2:**
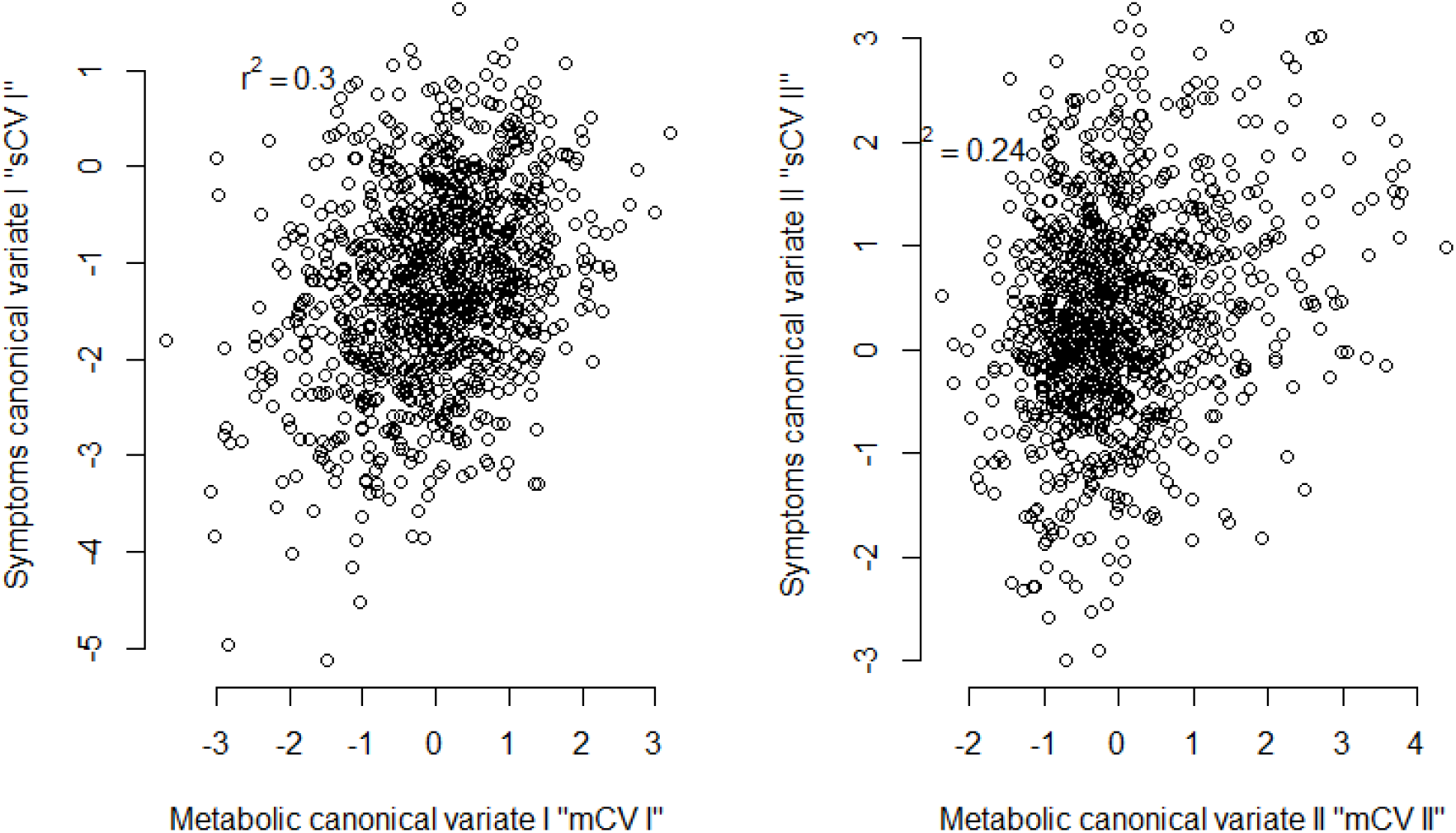
canonical correlation between first and second canonical pairs

**Supplemental table 2.**
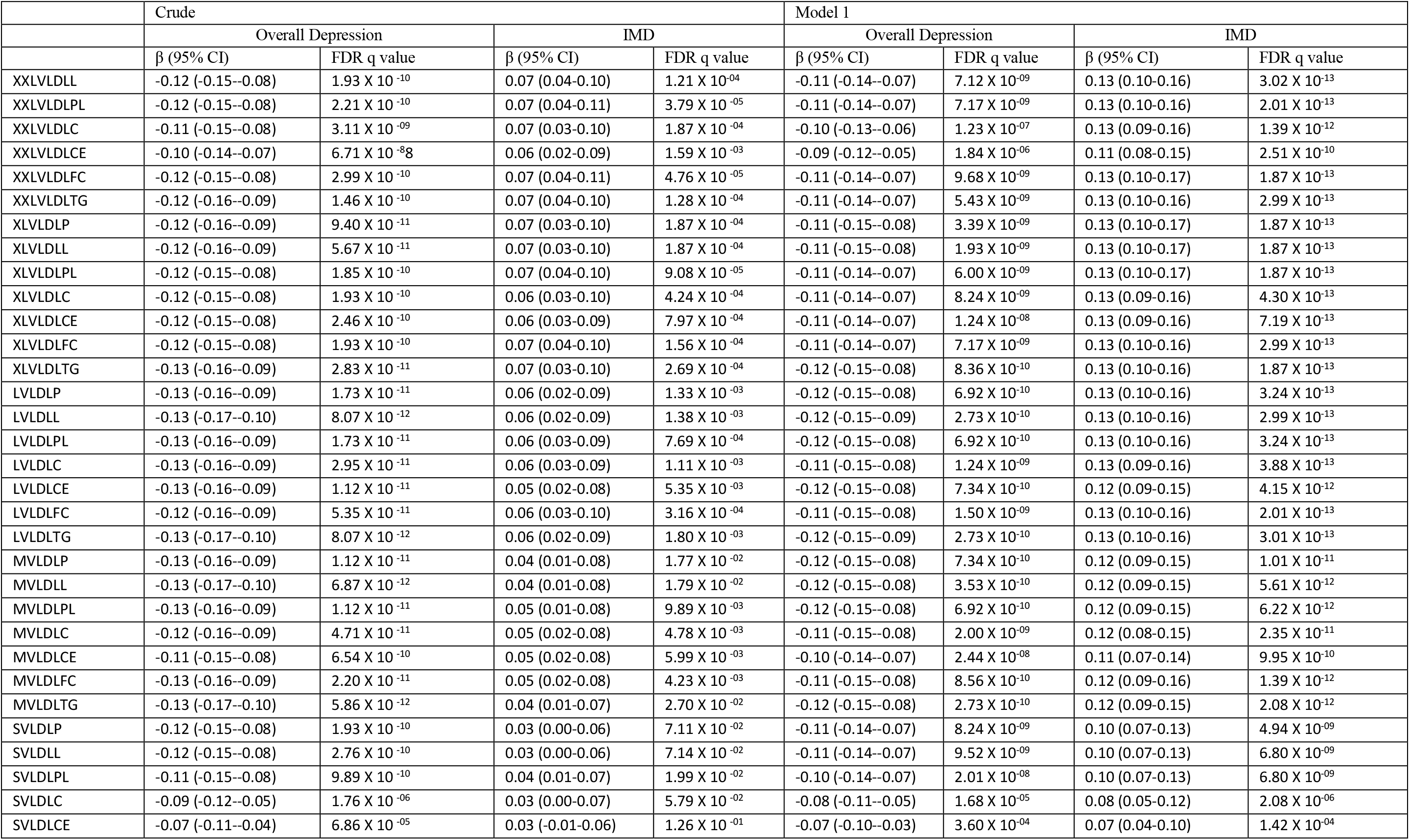
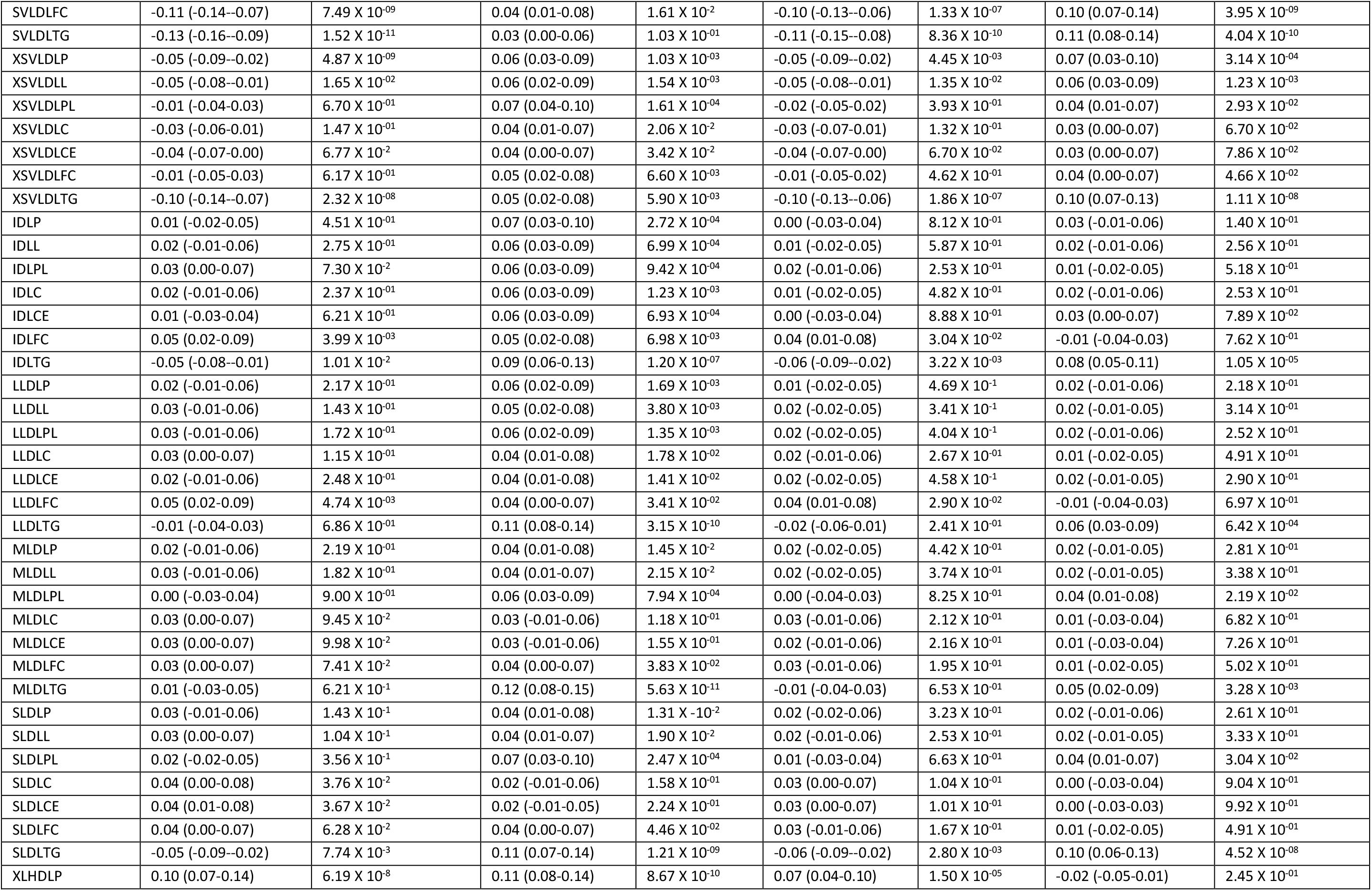
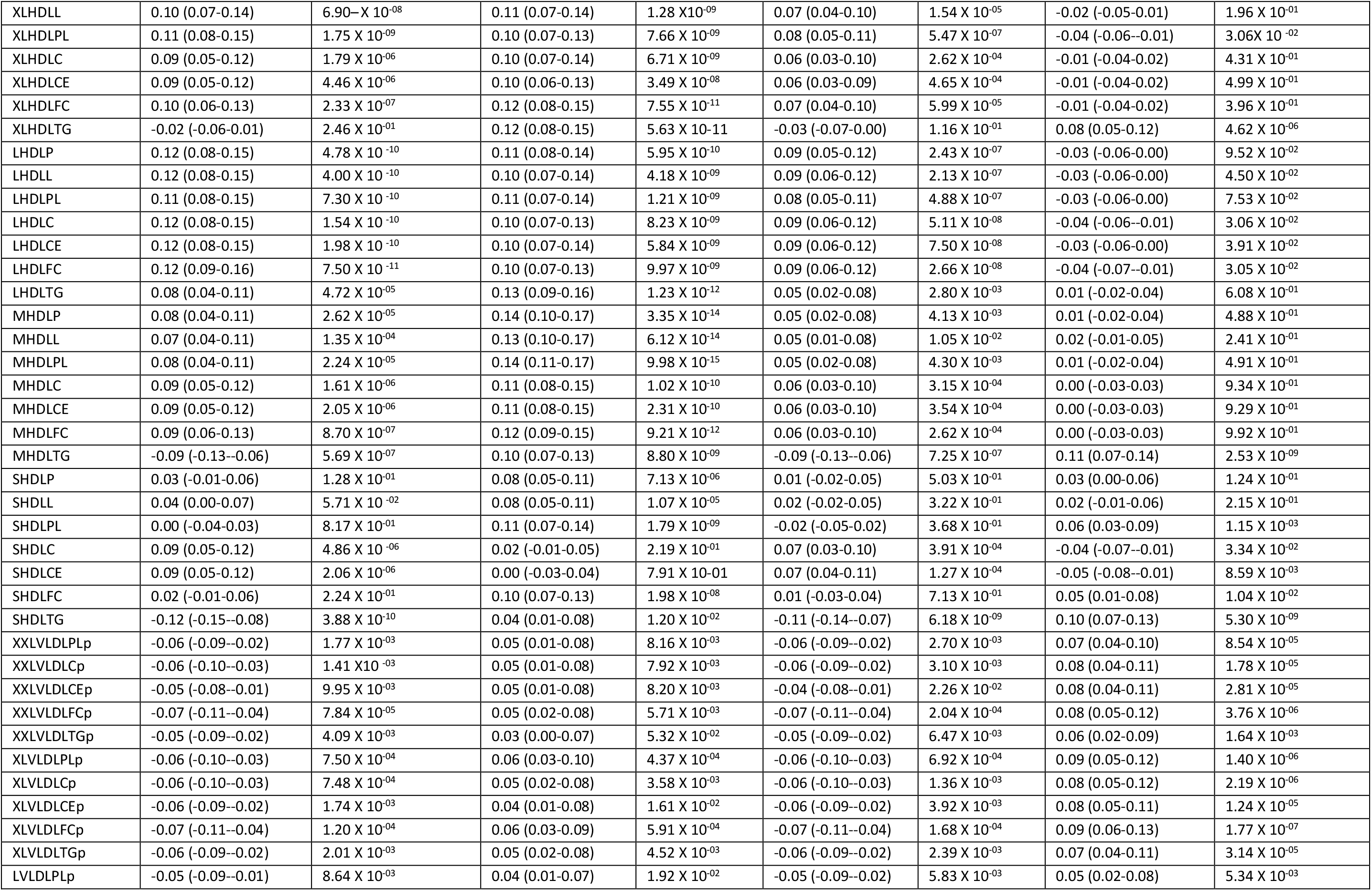
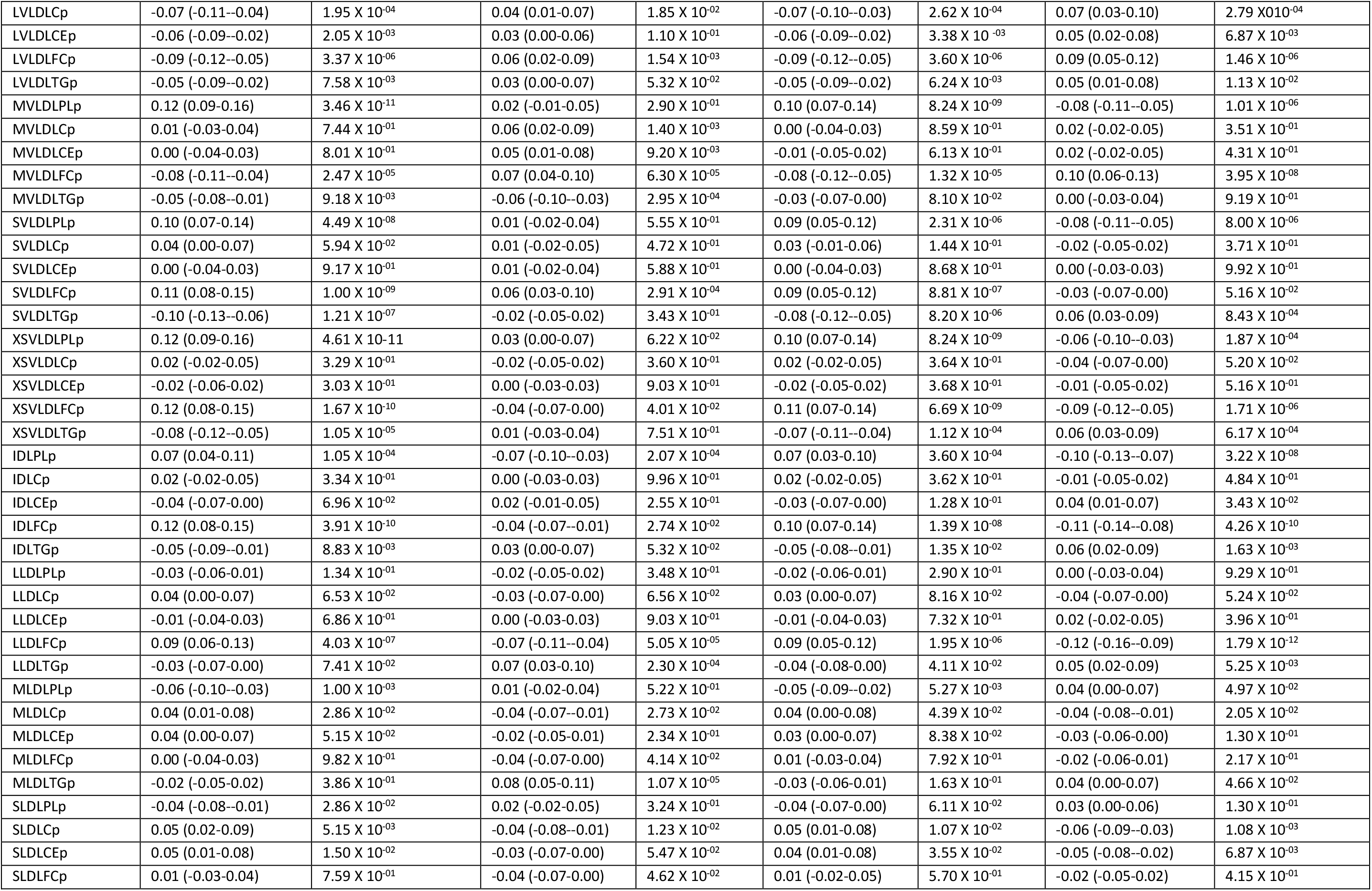
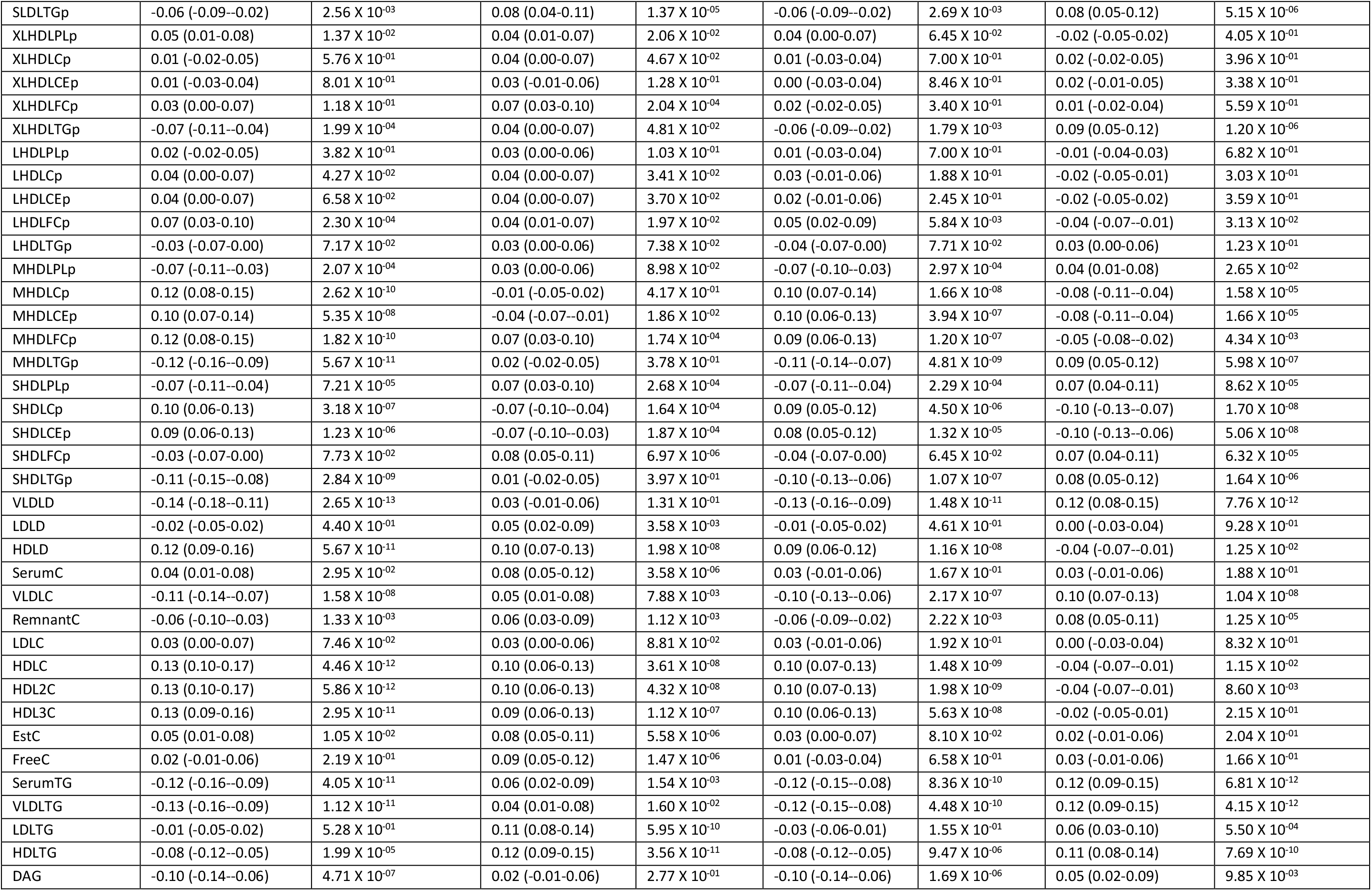
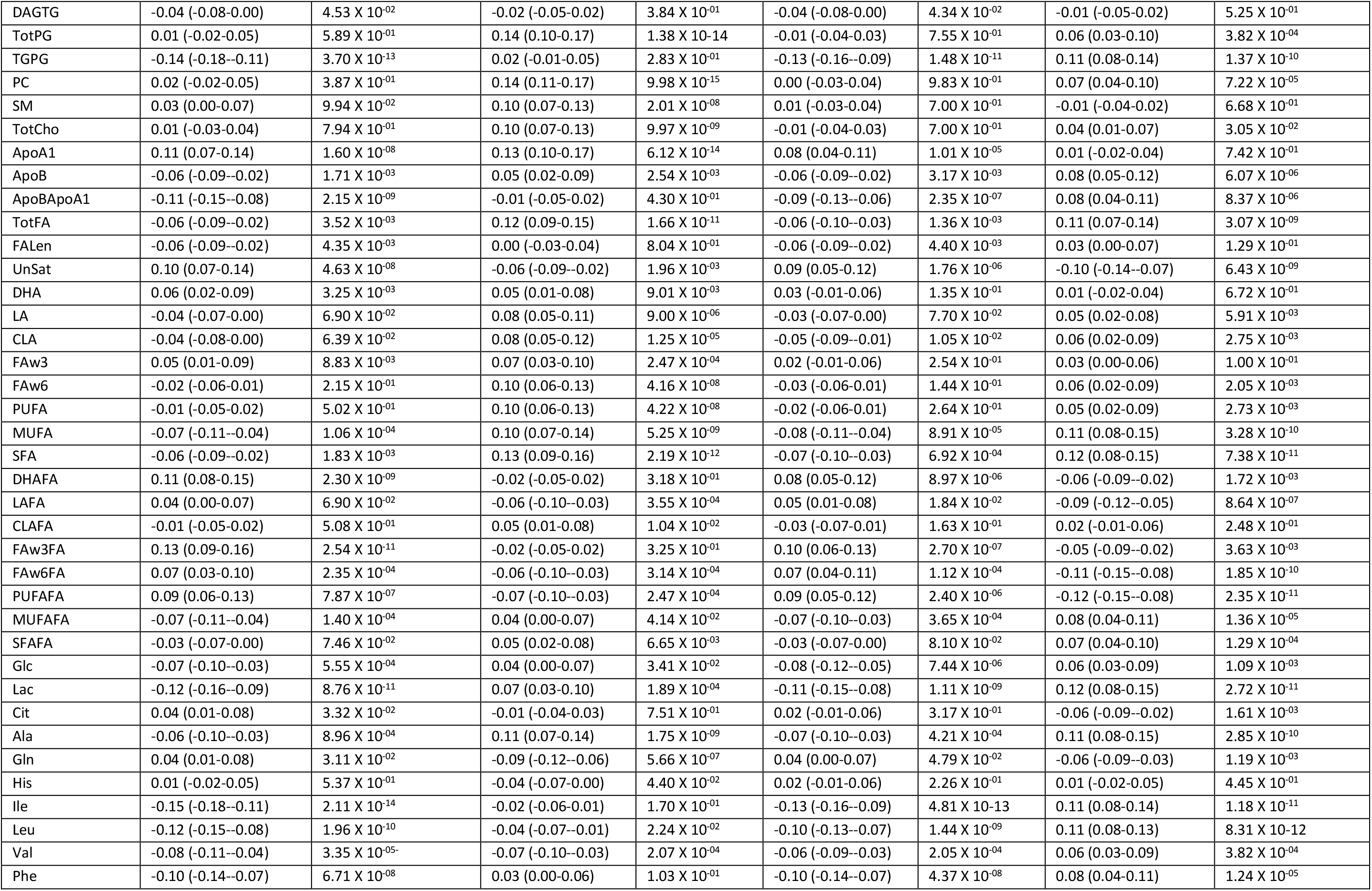
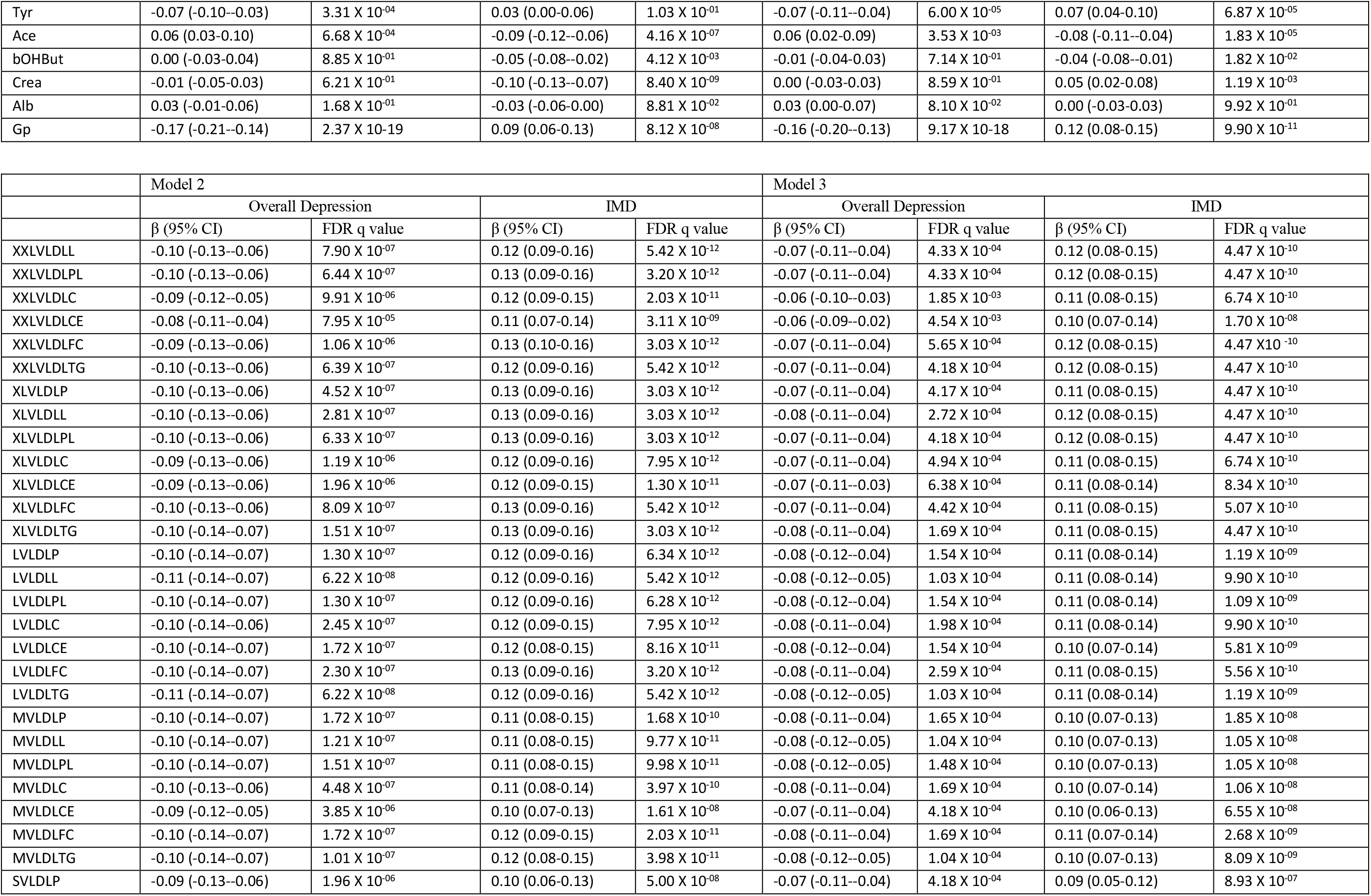
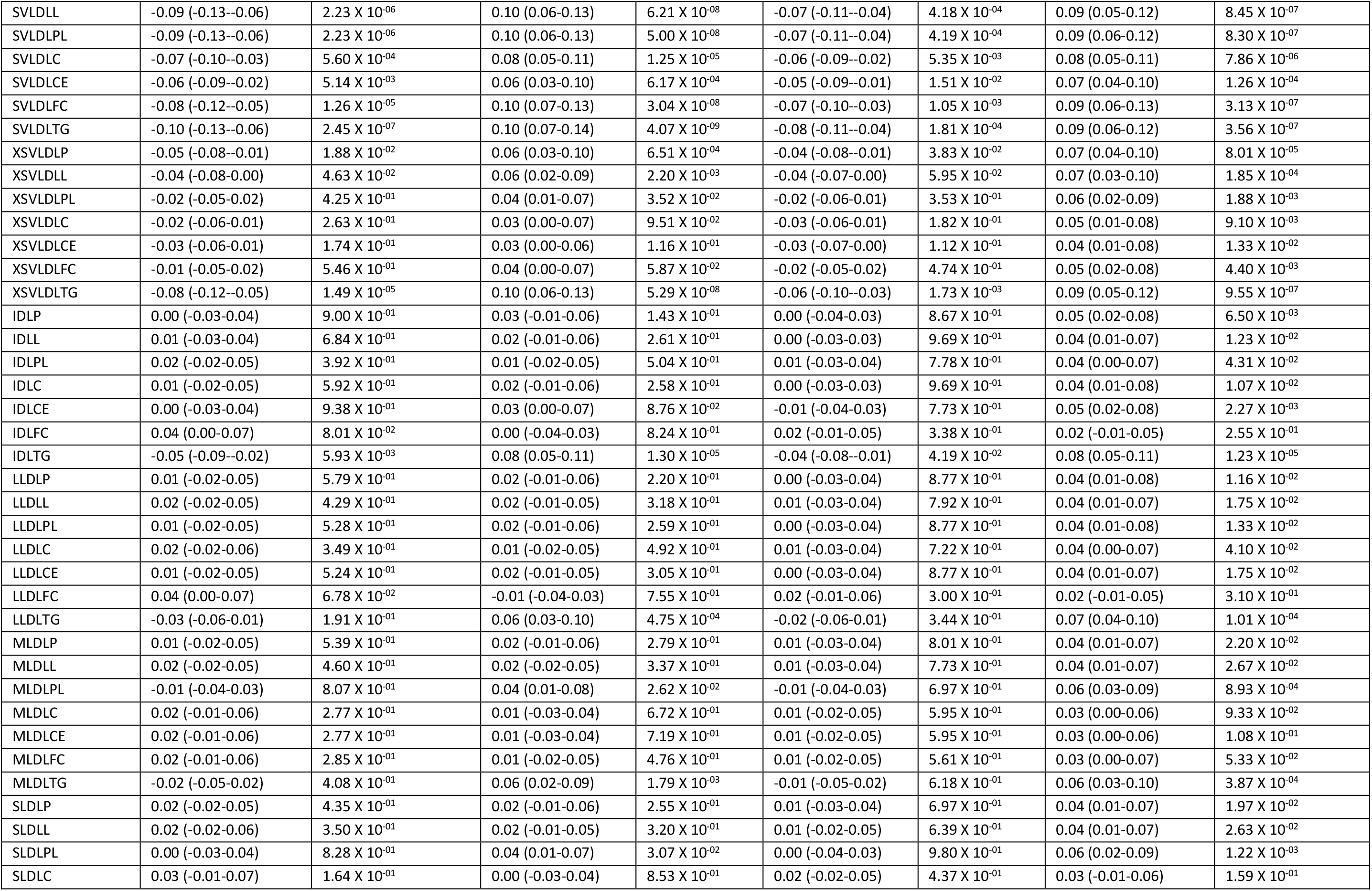
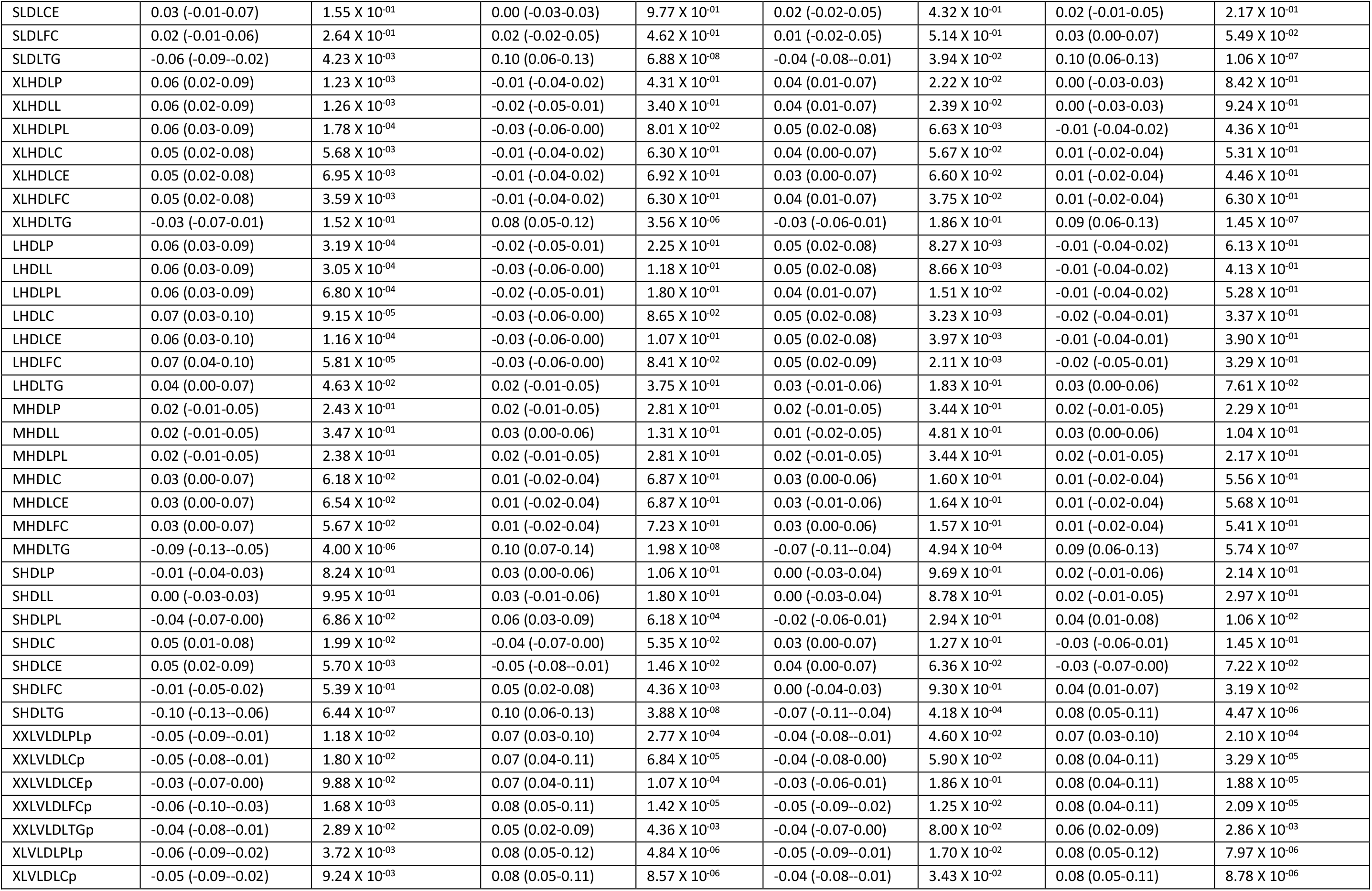
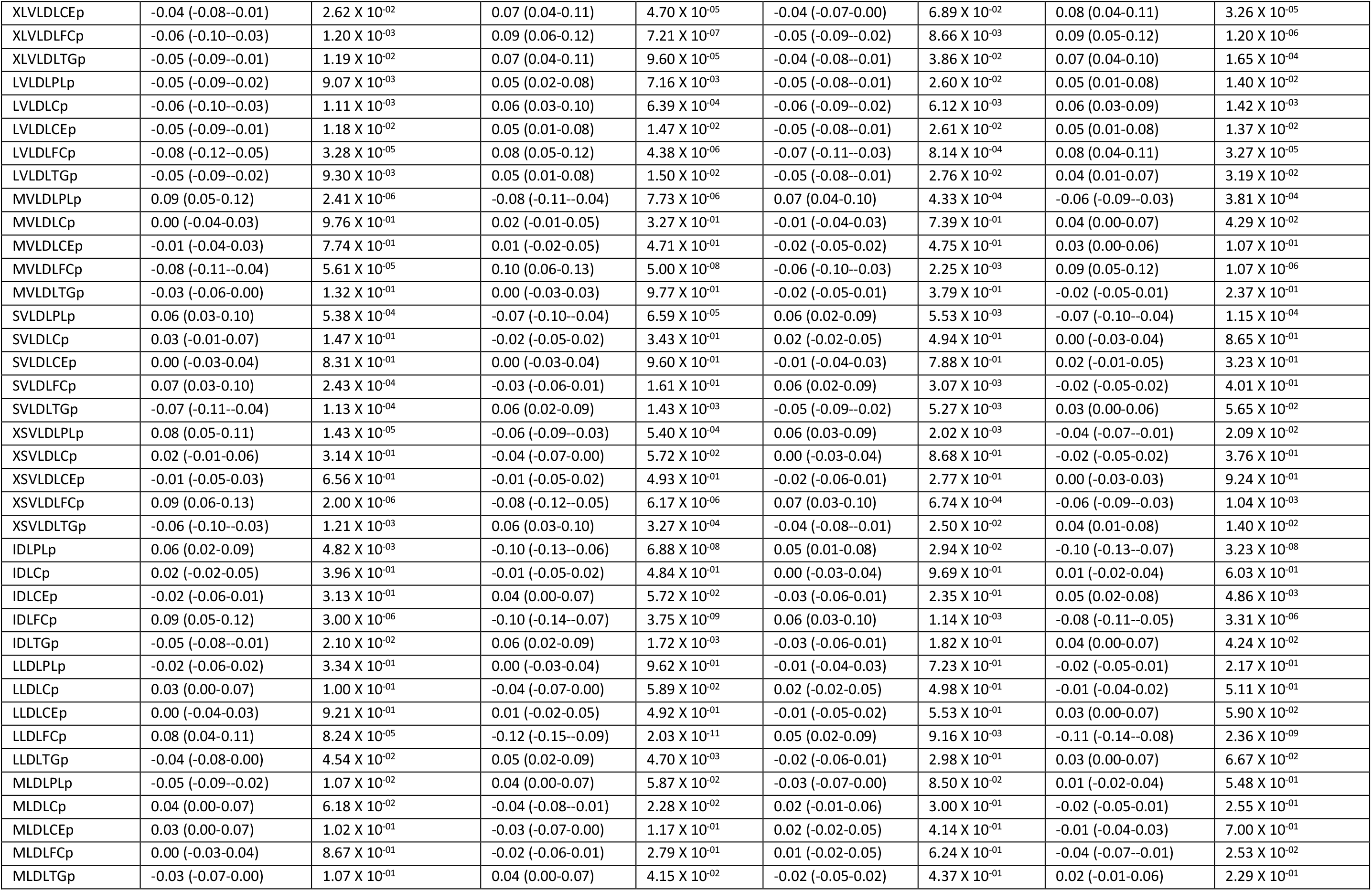
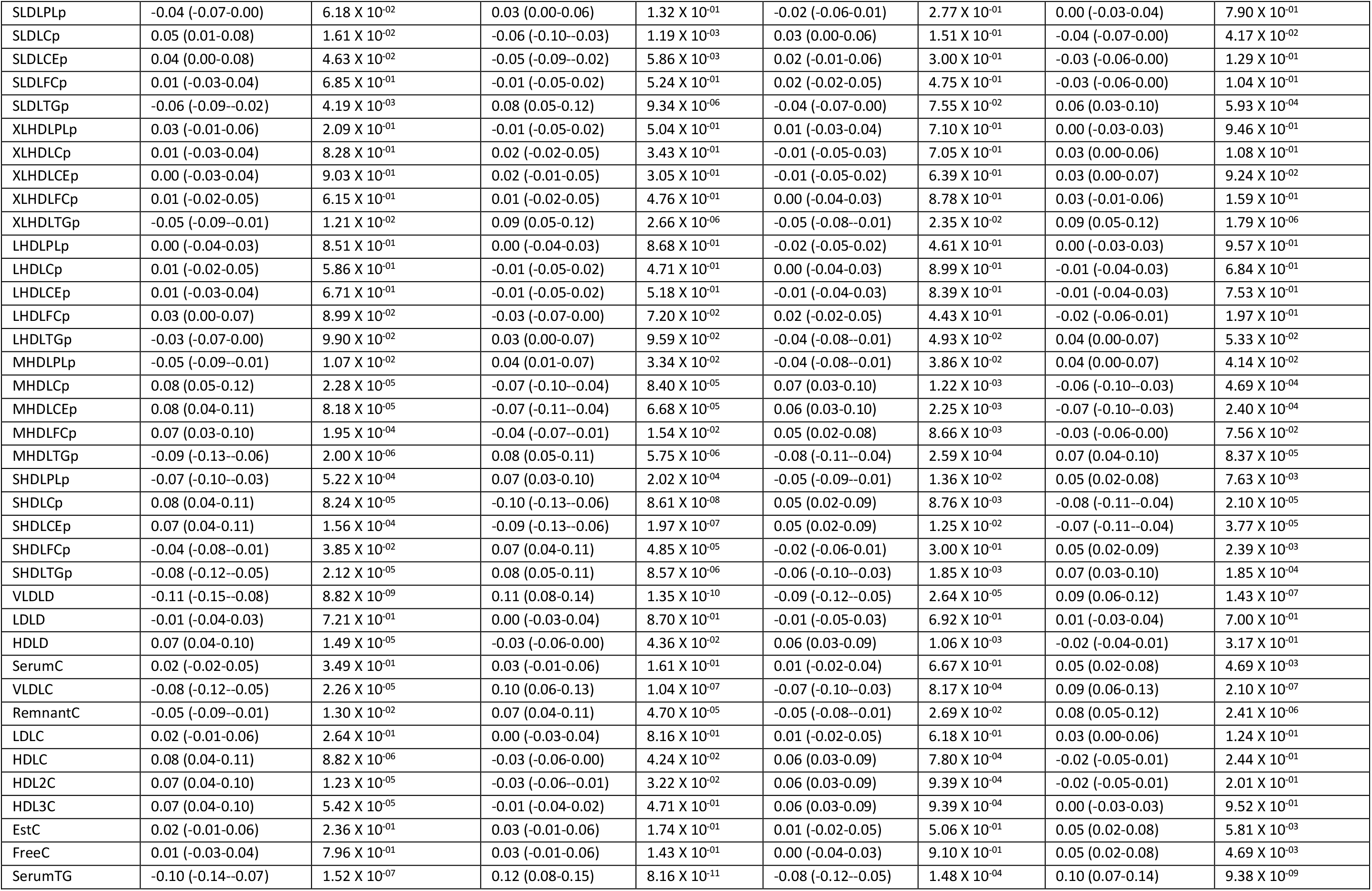
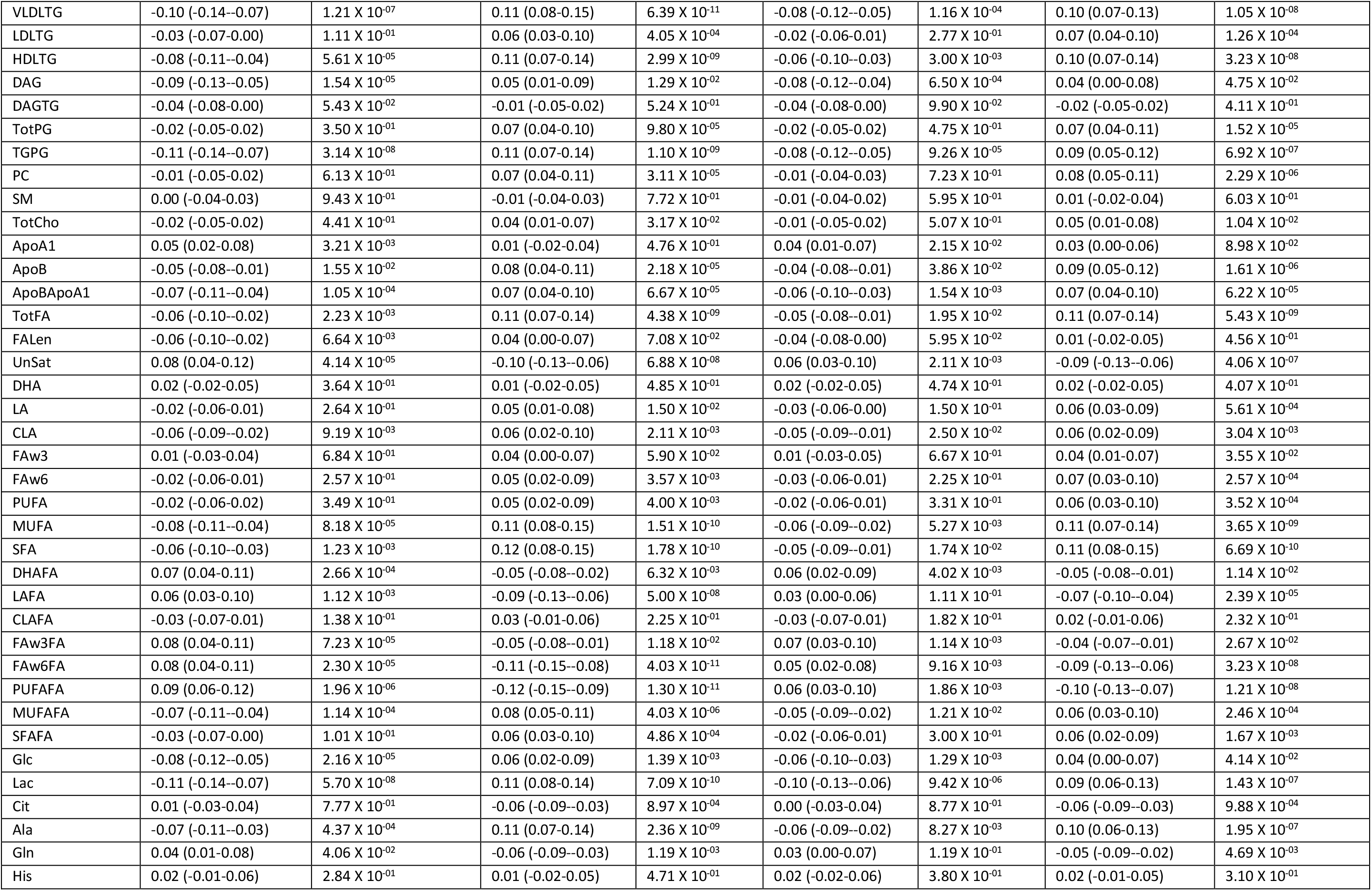
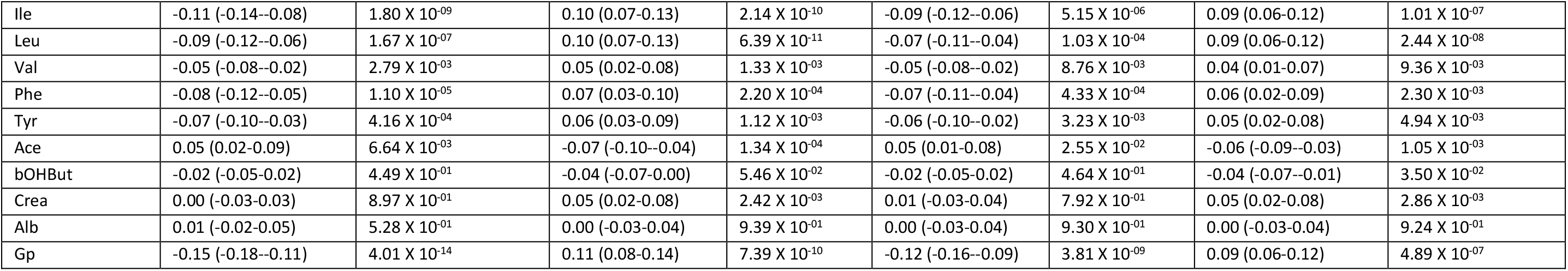
The linear regression analysis of the association between the sum score of the symptoms extracted from the metabolite-symptom clustering step and the metabolites.

**Supplemental table 3.** The linear regression analysis of the association between the sum score of the symptoms extracted from the metabolite-symptom clustering step and the cardiometabolic endpoints.

|  |  | Crude |  | Model 1 |  | Model 2 |  | Model 3 |  |
| --- | --- | --- | --- | --- | --- | --- | --- | --- | --- |
|  |  | Overall Depression | IMD | Overall Depression | IMD | Overall Depression | IMD | Overall Depression | IMD |
| Cardiometabolic endpoints | SD | β (95% CI) | β (95% CI) | β (95% CI) | β (95% CI) | β (95% CI) | β (95% CI) | β (95% CI) | β (95% CI) |
| BMI (kg/m<sup>2</sup>) | 4.43 | -0.17<br>(-0.20--0.13) | 0.13<br>(0.09-0.16) | -0.16<br>(-0.19--0.12) | 0.11<br>(0.08-0.15) | -0.15<br>(-0.19--0.12) | 0.11<br>(0.07-0.14) | -0.13<br>(-0.17--0.09) | 0.10<br>(0.06-0.13) |
| Total body fat (%) | 8.62 | -0.08<br>(-0.12--0.05) | 0.30<br>(0.27-0.33) | -0.11<br>(-0.13--0.09) | 0.09<br>(0.06-0.11) | -0.11<br>(-0.13--0.09) | 0.08<br>(0.06-0.11) | -0.09<br>(-0.12--0.07) | 0.07<br>(0.05-0.10) |
| Waist circumference (cm) | 13.34 | -0.17<br>(-0.20--0.13) | 0.04<br>(0.01-0.07) | -0.16<br>(-0.19--0.12) | 0.12<br>(0.09-0.15) | -0.15<br>(-0.18--0.12) | 0.11<br>(0.08-0.15) | -0.13<br>(-0.16--0.09) | 0.10<br>(0.07-0.13) |
| Visceral adipose tissue (cm<sup>2</sup>)* | 13.34 | -0.17<br>(-0.23--0.11) | 0.02<br>(-0.03-0.08) | -0.15<br>(-0.20--0.10) | 0.11<br>(0.06-0.16) | -0.14<br>(-0.20--0.09) | 0.09<br>(0.04-0.14) | -0.12<br>(-0.18--0.07) | 0.07<br>(0.02-0.12) |
| Fasting glucose (mmol/L) | 0.96 | -0.07<br>(-0.10--0.03) | 0.06<br>(0.03-0.10) | -0.08<br>(-0.11--0.04) | 0.08<br>(0.05-0.11) | -0.07<br>(-0.11--0.04) | 0.08<br>(0.05-0.11) | -0.05<br>(-0.09--0.02) | 0.06<br>(0.03-0.09) |
| HOMA-1B | 67.52 | -0.11<br>(-0.14--0.07) | 0.06<br>(0.03-0.10) | -0.09<br>(-0.13--0.06) | 0.08<br>(0.05-0.12) | -0.08<br>(-0.12--0.05) | 0.08<br>(0.04-0.11) | -0.08<br>(-0.12--0.05) | 0.07<br>(0.04-0.11) |
| HOMA-IR | 0.96 | -0.10<br>(-0.14--0.07) | 0.08<br>(0.05-0.11) | -0.10<br>(-0.14--0.07) | 0.10<br>(0.06-0.13) | -0.10<br>(-0.13--0.06) | 0.09<br>(0.06-0.13) | -0.08<br>(-0.12--0.04) | 0.08<br>(0.05-0.12) |
| HbA1c (%) | 0.46 | -0.10<br>(-0.13--0.06) | 0.09<br>(0.06-0.13) | -0.11<br>(-0.15--0.08) | 0.09<br>(0.05-0.12) | -0.09<br>(-0.13--0.06) | 0.09<br>(0.05-0.12) | -0.08<br>(-0.11--0.04) | 0.07<br>(0.04-0.10) |
| Total cholesterol (mmol/L) | 1.05 | 0.02<br>(-0.02-0.05) | 0.08<br>(0.05-0.11) | 0.00<br>(-0.03-0.04) | 0.04<br>(0.01-0.07) | 0.00<br>(-0.04-0.03) | 0.04<br>(0.00-0.07) | -0.01<br>(-0.04-0.03) | 0.06<br>(0.03-0.09) |
| HDL-cholesterol (mmol/L) | 0.46 | 0.13<br>(0.10-0.17) | 0.09<br>(0.06-0.13) | 0.10<br>(0.07-0.13) | -0.04<br>(-0.07--0.01) | 0.07<br>(0.04-0.10) | -0.03<br>(-0.06-0.00) | 0.06<br>(0.03-0.09) | -0.02<br>(-0.05-0.01) |
| Triglycerides (mmol/L) | 0.84 | -0.13<br>(-0.17--0.10) | 0.06<br>(0.03-0.10) | -0.12<br>(-0.16--0.09) | 0.12<br>(0.09-0.15) | -0.11<br>(-0.14--0.07) | 0.12<br>(0.08-0.15) | -0.09<br>(-0.12--0.05) | 0.11<br>(0.07-0.14) |
| LDL-cholesterol (mmol/L) | 0.96 | 0.02<br>(-0.01-0.06) | 0.02<br>(-0.01-0.05) | 0.02<br>(-0.02-0.05) | 0.00<br>(-0.03-0.04) | 0.02<br>(-0.02-0.05) | 0.00<br>(-0.03-0.04) | 0.01<br>(-0.03-0.04) | 0.02<br>(-0.01-0.06) |

Number of individuals with data for BMI: 6572, Total body fat: 6541, Waist circumference: 6566, Visceral adipose tissue: 1869, Fasting glucose: 6554, HOMA-1B: 6541, HOMA-IR: 6545, HbA1c: 6543, Total cholesterol: 6562, HDL-cholesterol: 6561, Triglycerides: 6561, LDL-cholesterol: 6560. Model 1: adjusted for age, sex, and education. Model 2 adjusted for, age, sex, education, smoking, alcohol consumption, caloric intake, and physical activity. Model 3 adjusted for, age, sex, education, smoking, alcohol consumption, caloric intake, physical activity, lipid lowering drugs, and. antidepressants.

## Notes

### Author Declarations

The research protocol of NESDA was approved by the medical ethical committees of the following participating universities: Leiden University Medical Center (LUMC), Vrije University Medical Center (VUMC), and University Medical Center Groningen (UMCG). The NEO study was approved by medical ethics committee of Leiden University Medical Center (LUMC). All participants gave written informed consent.

